# A predictive model of ebolavirus spillover incorporating change in forests and human populations across spatial and temporal scales

**DOI:** 10.1101/2023.08.29.23294795

**Authors:** Carson T. Telford, Brian R. Amman, Jonathan S. Towner, Sarah Bowden, Joel M. Montgomery, Justin Lessler, Trevor Shoemaker

**Affiliations:** Gillings School of Global Public Health, University of North Carolina, Chapel Hill, North Carolina; Viral Special Pathogens Branch, Centers for Disease Control and Prevention, Atlanta, Georgia; Division of Global Migration and Quarantine, Centers for Disease Control and Prevention, Atlanta, Georgia; Carolina Population Center, University of North Carolina, Chapel Hill, North Carolina; Bloomberg School of Public Health, Johns Hopkins University, Baltimore, Maryland

## Abstract

Past research has found associations between ebolavirus spillover and forest loss and fragmentation, although most predictions of the spatial distribution of ebolaviruses have not utilized these data. Spatial and temporal scales of covariate data measurement have also not previously been accounted for in predictive models of ebolavirus spillover, making it difficult to account for variables that influence ebolavirus transmission dynamics, such as movement and interaction of human and animal populations, trade, and animal behavioral responses to human presence and a changing environment. Using annual data on forest cover, forest loss and fragmentation, human population distribution, and meteorological variables, we developed models of ebolavirus spillover events from 2001-2021 to estimate the annual relative odds of ebolavirus spillover in equatorial Africa in 2021 and 2022. Analyses were done separately for all ebolavirus species (All-species analysis) and Zaire ebolavirus alone (Zaire-only analysis). Locations with the highest estimated relative odds of spillover occurred in patches throughout Democratic Republic of the Congo (DRC), Republic of the Congo, Gabon, Cameroon, and coastal west Africa in a spatial trend that resembled that of maps of forest fragmentation and forest loss. Reduced analyses that ignored forest loss and fragmentation data produced estimates that were distinct from the full analyses, highlighting locations where forest loss and fragmentation drove model predictions, which included coastal west Africa, southern Cameroon, and parts of DRC. Estimated spillover odds and increase in spillover odds at 2022 spillover sites ranked among the highest in equatorial Africa, suggesting the potential of predictive analyses to prioritize locations for surveillance and research efforts.

**Significance:** Using annually updated data on forest change, human population distribution, and meteorological conditions, we developed models to estimate the relative odds of ebolavirus spillover in 2021 and 2022 in equatorial Africa, identifying locations with elevated odds of spillover and temporal shifts in spillover odds between the two years. Locations were identified whose estimates of spillover odds were driven by forest change. During 2022, two ebolavirus spillover events were identified in locations with elevated spillover odds estimates. Annually updated estimates of ebolavirus spillover can guide public health surveillance programs to high-risk locations at specific times, and guide potential ebolavirus reservoir sampling efforts to locations that recently experienced increases in spillover odds.

## Introduction

Outbreaks of disease caused by ebolaviruses usually result from zoonotic spillover into a susceptible human from an infected animal, followed by secondary transmission in humans (1–4). Ebolavirus spillover into humans is rare, though the rate of identification of ebolavirus spillover events is increasing, likely due to both increased surveillance capacity in affected countries and increased intersection between humans and animals (5,6). Over the 23-year span from 1976-1999, ebolavirus spillover was only identified 10 times: a rate of less than one every two years. In the most recent 23 years from 2000-2023, 25 spillover events have been identified: a rate of more than one per year (7). Recent outbreaks have also demonstrated the large-scale public health threat presented by these viruses, most notably the west African outbreak in Guinea, Sierra Leone, and Liberia in 2014-2016 which caused nearly 30,000 cases and over 11,000 deaths, and the Democratic Republic of the Congo (DRC) outbreak from 2018-2020 which caused approximately 3,500 cases and 2,300 deaths (8).

Four species of ebolavirus have been identified that can cause human disease (Zaire, Sudan, Bundibugyo, and Tai Forest ebolaviruses), and spillover of these viruses has occurred in heavily forested regions throughout equatorial Africa. The natural host(s) of ebolaviruses remain unknown, though evidence suggests that African forest-dwelling bats are likely natural reservoirs (3,9). Due to differing ecological and geographic contexts associated with spillover of each ebolavirus species, it has been suggested that each ebolavirus species may have a unique reservoir animal species (10,11). A similar “one virus-one host species” relationship was suggested following a study of inoculation of filoviruses in *Rousettus aegyptiacus* bats (a confirmed reservoir of Marburg virus), which determined that these bats were only able to host and transmit one type of filovirus (12).

Multiple efforts have been made to estimate the spatial distribution of the ebolavirus habitat suitability and spillover density using data on vegetation density, meteorology, and bat distribution, all of which suggest the locations with highest likelihood of ebolavirus presence and spillover are spread across equatorial Africa in countries such as the Democratic Republic of the Congo (DRC), the Republic of the Congo (ROC), Gabon, Cameroon, Guinea, and Uganda (11,13–16). Although underlying mechanism is unclear, associations have been found between ebolavirus spillover and forest loss and fragmentation, or the division of contiguous forests into smaller patches often due to anthropogenic land conversion such as road construction, agricultural expansion, and urban development (17,18).

Past spillover of ebolaviruses has been linked to contact with infected animals such as apes and duikers, hunting, and proximity to bats, and human-animal contact rates may increase as human population sizes increase and expand into previously undisturbed animal habitats (17–19). However, different animal species may have varying responses to habitat destruction, potentially resulting in different levels of spillover risk across ebolavirus species. For example, some bat species may be displaced or forced to travel greater nightly distances to forage due to resource scarcity, which could induce sufficient immune stress to cause viral shedding (5,20–22), while others may adapt to habitat disruption by residing near human settlements, especially if deforested areas are used to cultivate fruit crops that bats may forage, potentially increasing contact between humans and bats and the possibility of spillover (23–25). Changes in the spatial distribution of human populations, especially at forest edges, can increase the population of humans at risk of infection and is often directly associated with forest loss (26,27).

While forested areas with increasing levels of forest loss, fragmentation, and human populations may be at elevated risk of ebolavirus spillover, the spatial and temporal scales at which these variables influence and are predictive of spillover risk is unclear due to the variation in movement patterns among individuals associated with transmission patterns. For example, the spatial scale of human population distribution may vary in its association with spillover risk, as bushmeat hunting can occur in forested areas surrounding human settlements, and trade of animal products can occur between villages and markets (28). Among suspect ebolavirus reservoirs, nightly flight ranges of forest dwelling bat species range from 1 to >50 kilometers (29–31), while migratory species have been observed to travel up to 300 kilometers in a single night, and 2,500 kilometers over a 5-month span (32). Movement patterns of susceptible animal species such as non-human primates may also vary in response to forest loss and increased presence of human activities (33,34). Temporal lags may also exist between human encroachment into forests and zoonotic spillover depending on the response of ebolavirus reservoirs to such habitat disruption.

Given these complex relationships between potential predictors of ebolavirus spillover risk across various spatial and temporal scales of measurement, we sought to develop a model of ebolavirus spillover events from 2001-2021 that could be used to estimate the annual relative likelihood of ebolavirus emergence in 2021 and 2022 across equatorial Africa, incorporating forest cover, forest loss and fragmentation, and human population data across varying spatial and temporal scales in combination with other relevant environmental predictors. Estimated spillover odds from models incorporating forest change data were compared to models ignoring forest change to identify locations where estimates were driven by forest change information. Temporal shifts in spillover odds due to changes in the environment and human population sizes were evaluated from 2021-2022, and relative spillover odds estimates were also evaluated at the locations of two ebolavirus spillover events that occurred in 2022.

## Results

### Descriptive analysis

Forest cover, loss, and fragmentation were measured using non-overlapping circular “donut” buffers of 10-, 25-, 50-, 100-, and 150-kilometer radii surrounding 22 ebolavirus spillover locations from 2001-2021 and compared to absence locations where ebolavirus emergence was not detected. Forest loss was measured during the same (FL SY), one year prior (FL 1YP), and two years prior (FL 2YP) to spillover events. Across the spatial scales of measurement, average forest cover surrounding spillover sites of all ebolavirus species combined ranged from 69.5-76.1% while it ranged from 37.9-38.6% around absence locations. Average annual forest loss surrounding spillover sites ranged from 0.23-0.66% and surrounding absence locations ranged from 0.14-0.16% across spatial and temporal scales of measurement. The average proportion of fragmented forest area surrounding spillover sites ranged from 26.3-30.6% and surrounding absence sites ranged from 11.9-12.4% across spatial scales. Annual forest loss percentage was consistently highest when measured within 10-kilometers of spillover sites (FL SY: 0.50%; FL 1YP: 0.53%; FL 2YP: 0.66%). Average log human population count was lower on average surrounding spillover sites (9.2-13.3) compared to absence locations (9.3-14.3). However, the product term between log population count and forest cover was higher surrounding spillover sites. Historical average variables including PET, elevation, precipitation seasonality, and temperature seasonality tended to be lower surrounding spillover sites compared to absence locations, whereas NTLST was slightly higher (Supplemental Table 3).

Compared to the environmental conditions averaged across all ebolavirus species (species-averaged), Zaire ebolavirus spillover sites tended to have higher proportions of forest cover, forest loss, and forest fragmentation across all spatial and temporal scales, and lower log human population count across all spatial scales. The number of Sudan and Bundibugyo virus spillover events was too small to make species-stratified comparisons.

### Predictive model results

Models were fit using boosted regression trees (BRT) to estimate the odds of ebolavirus spillover, and 3-fold cross-validation was used to evaluate predictive accuracy, where the area under the receiver operator curve (AUC) for the All-species analysis was 0.88, and for the Zaire-only analysis was 0.91 (Figure 1). Reduced versions of each analysis were also conducted excluding forest loss and fragmentation data and resulted in slightly higher cross validation AUC for the All-species analysis (AUC=0.90), and lower cross validation AUC for the Zaire-only analysis (AUC=0.90). To classify spillover and non-spillover locations, we used the fitted spillover odds cutoff that that maximized the product between sensitivity and specificity, which resulted in sensitivity of 86.4% and specificity of 72.7% in the All-species full model, sensitivity of 95.5% and specificity of 72.8% in the All-species reduced model, sensitivity of 93.8% and specificity of 86.6% in the Zaire-only full model, and sensitivity of 87.5% and specificity of 91.6% in the Zaire-only reduced model (Figure 1).

**Figure 1.**
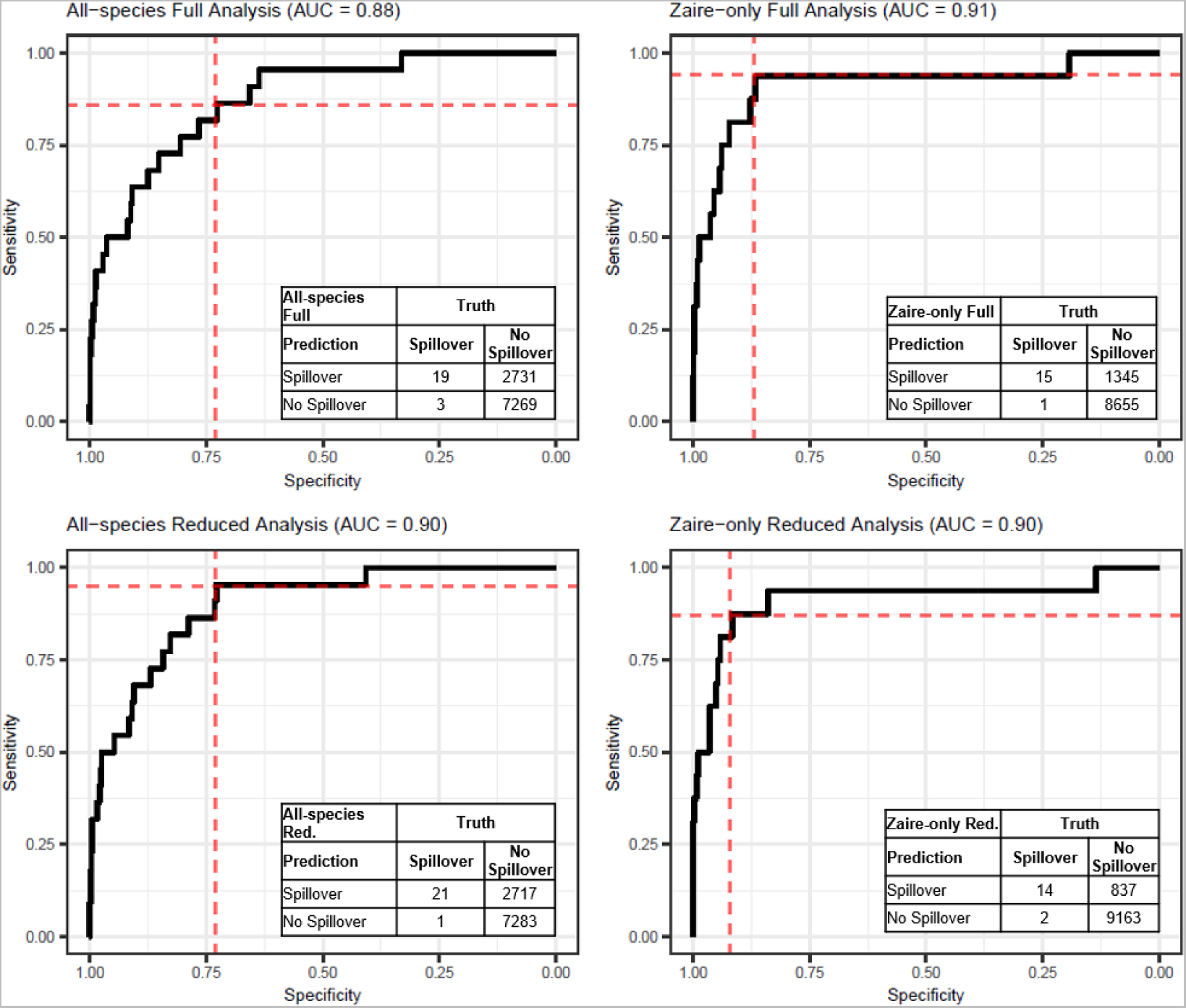
Receiver operator curves (ROC) visualizing sensitivity and specificity in predicting ebolavirus spillover from the All-species full and reduced analyses (left) and Zaire-only full and reduced analyses (right) based on 3-fold cross validation and corresponding confusion matrices resulting from each analysis.

In the All-species analysis, log human population at spatial scales of 50-100 kilometers and <10 kilometers were among covariates with the highest median relative importance in predicting spillover. Interestingly when measuring human population size 50-100 kilometers from spillover sites, marginal response curves showed spillover likelihood decreased as human populations increased. However, when measured within 10 kilometers, spillover likelihood increased as human population size increased. The product term between log population count and forest cover percentage had the third highest median relative prediction importance and had a positive relationship with spillover likelihood, suggesting that spillover likelihood was highest in locations with both large populations and high percentages of forest cover. Forest fragmentation had the highest relative importance in spillover prediction when measured at a spatial scale of 100-150 kilometers surrounding spillover sites, whereas the forest loss variables with the highest relative predictive importance were forest loss <10 kilometers from spillover sites during the same year and 2 years prior to spillover events. Marginal response curves showed that spillover likelihood increased as forest loss and fragmentation increased across all spatial and temporal scales of analysis. Precipitation seasonality was also among the most important predictors of spillover in the All-species analysis, and spillover likelihood tended to decrease as annual seasonality of monthly precipitation increased (Figure 2).

**Figure 2.**
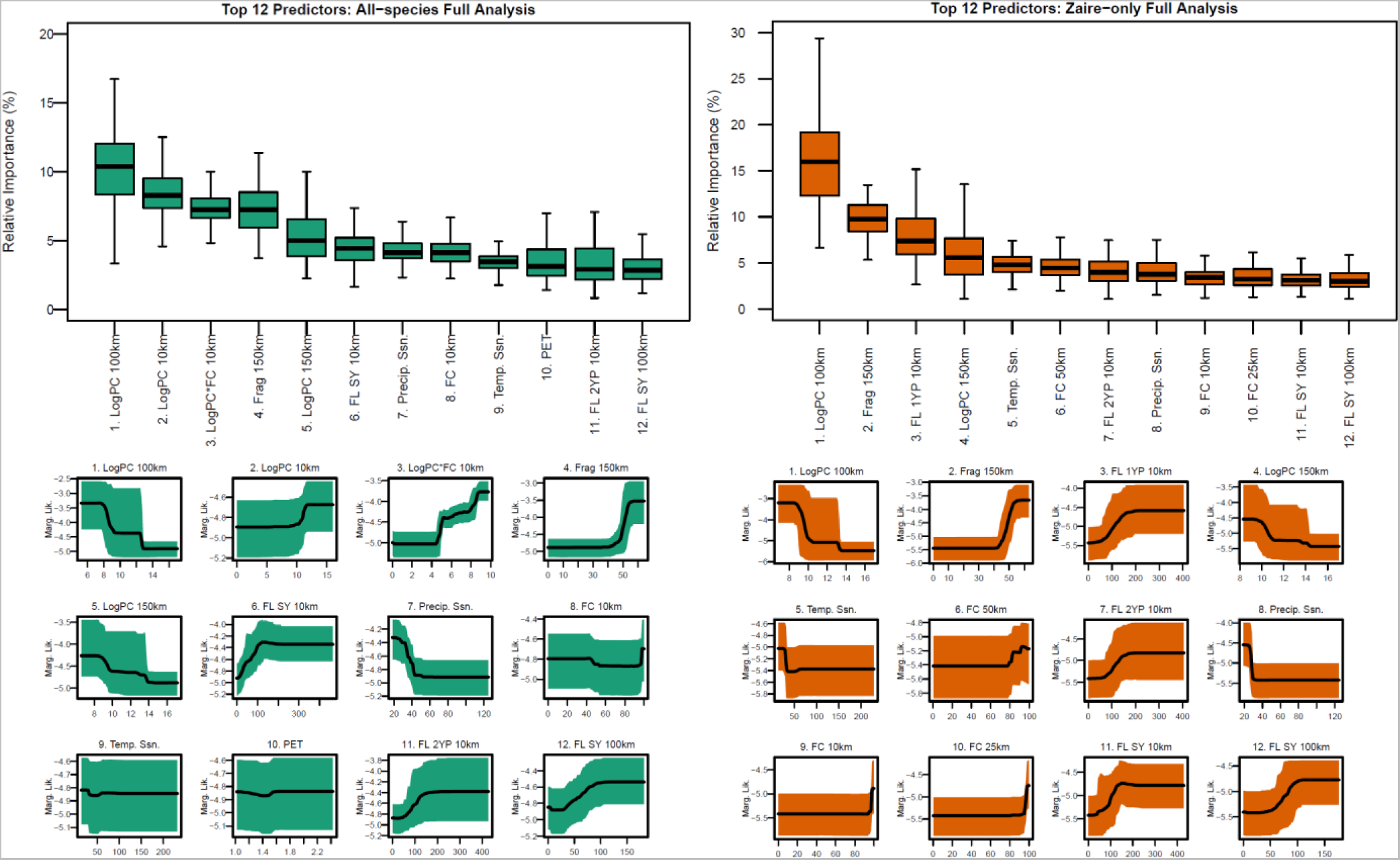
Relative variable importance in predicting ebolavirus spillover (top) and marginal response curves of the relationship between spillover likelihood and each covariate (bottom) for the top 16 predictors in the All-species analysis (left) and Zaire-only analysis (right). Abbreviations: PC=Population count; FC=Forest Cover; Frag=Fragmentation; FL=Forest Loss; SY=Same Year; Precip=Precipitation; Temp=Temperature; Ssn=Seasonality; PET=Potential Evapotranspiration; 1YP=One year prior; 2YP=Two years prior; km=kilometers;

In the Zaire-only analysis, relative importance and marginal response curves of covariates were similar to those from the All-species analysis with some exceptions. While log population count 50-100 kilometers from spillover sites remained the variable with the highest relative predictive importance, median relative importance across the model ensemble increased from 10% in the All-species analysis to 16% in the Zaire-only analysis, where spillover likelihood was highest where population size was low. Variables related to forest loss and fragmentation had higher relative importance in the Zaire-only analysis compared to the All-species analysis, including forest fragmentation 100-150 kilometeres from spillover sites, forest loss within 10 kilometers and during the same and prior 2 years to spillover events, and forest cover 25-50 kilometers from spillover sites. Positive associations were found between spillover likelihood and forest loss, forest cover, and fragmentation regardless of the spatial or temporal scale.

### Relative spillover odds estimation

Using the models trained on spillover events from 2001-2021, we estimated the odds of ebolavirus spillover, based on observed covariate values across the study area in the years 2021 and 2022 (the most recently available forest cover, forest loss, and human population data). Odds estimates were then converted into relative odds ratios (ROR) by dividing the estimated odds of spillover in each location by the overall mean estimated odds across the entire study area. Estimated spillover RORs >1 represented locations with above average odds of spillover. Estimated 2022 spillover RORs from the All-species analysis ranged from 0.3-32.3, and 21.0% of the study area had above-average spillover odds. In the Zaire-only analysis, estimated spillover RORs in 2022 ranged from 0.2-31.2 and 19.0% of the study area had above-average spillover odds (Figure 3).

**Figure 3.**
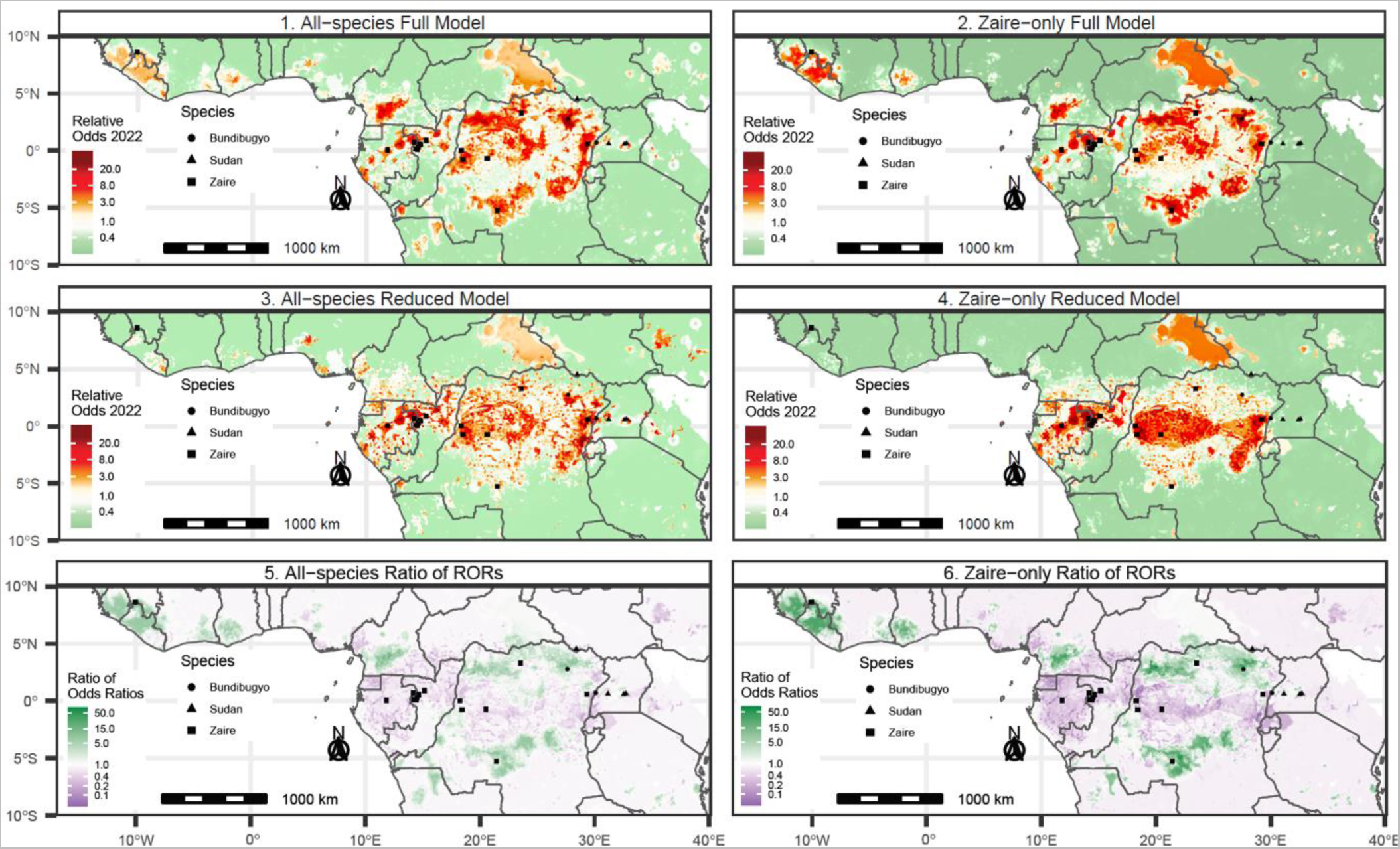
Model predictions of the relative odds of ebolavirus spillover in 2022 resulting from the All-species and Zaire-only analyses. The top row represents the estimated relative odds of ebolavirus spillover in the year 2022 from the full models, which included covariates representing forest loss and fragmentation across varying spatial and temporal scales. Maps in the second row represent estimated spillover odds resulting from models that did not include information on forest loss and forest fragmentation. The third row represents the ratio between estimated spillover odds resulting from the full and reduced versions of the All-species (left column) and Zaire-only (right column) analyses.

In the All-species analysis, six countries had an overall average spillover ROR >1, including DRC (2.0), Gabon (1.7), Liberia (1.7), Sierra Leone (1.2), Cameroon (1.1), and ROC (1.1). Seven countries also had at least one location with a spillover ROR >10, including DRC (max: 32.3), ROC (max: 28.1), Gabon (max: 25.9), Cameroon (max: 20.8), Uganda (max: 13.2), Equatorial Guinea (max: 12.5), and Central African Republic (max: 10.2). In the Zaire-only analysis, six countries had overall average spillover ROR >1, including Liberia (2.6), DRC (2.1), Sierra Leone (2.1), Gabon (1.8), Central African Republic (1.4), and ROC (1.1). Five countries had at least one location with an estimated Zaire virus spillover RORs >10, including DRC (max: 31.2), ROC (max: 23.6), Gabon (max: 23.6), Cameroon (max: 23.4), and Central African Republic (max: 10.3). The majority of 1x1 kilometer grid cells in the study area with an estimated ROR >1 occurred in DRC in the All-species (54.1%) and Zaire-only (58.1%) analyses. Areas outside and along the edges of dense forests tended to have higher spillover RORs in the All-species analysis compared to the Zaire-only analysis, whereas the highest estimates of spillover odds from the Zaire-only analysis occurred in areas with higher proportions of forest cover. Comparing estimates from the All-species and Zaire-only analyses, the areas with the greatest contrast in estimated spillover ROR occurred in Central Uganda and along the border area between Uganda, DRC, and Rwanda, where estimated spillover RORs from the All-species analysis were as much as 53.2 times and 24.9 times those from the Zaire-only analysis in Uganda and DRC, respectively. Prediction grid cells were also occurred in Nigeria, Kenya, Burundi, Rwanda, Angola, Cameroon, and Ethiopia where estimates from the All-species analysis were >10 times as high as those from the Zaire-only analysis (Supplemental Figure 2, Supplemental Table 4). In contrast, the Zaire-only analysis produced higher spillover ROR estimates throughout DRC, Sierra Leone, Liberia, along the border between Gabon and ROC, and in eastern Central African Republic. The largest contrast in estimated spillover odds from the Zaire-only analysis compared to the All-species analysis occurred in an area in DRC where spillover odds were as much as 5.1 times as high, and estimated RORs in the Zaire-only analysis reached >3 times as high as those from the All-species analysis in ROC, Gabon, Cote d’Ivoire, Liberia, Sierra Leone, and Ghana (Supplemental Table 4).

### Difference in Predictions between the Full and Reduced models

Compared to reduced models that excluded information on forest loss and fragmentation, incorporating these variables into models produced considerable differences in the spatial pattern of estimated spillover RORs across equatorial Africa, especially for estimates specific to Zaire ebolavirus. Whereas the highest estimates of spillover odds in the reduced version of the Zaire-only analysis were concentrated in a band along the equator primarily in the center of the rainforest DRC, ROC, and Gabon, estimates from the full model accounting for forest loss and fragmentation highlight patches away from the center of the rainforest where forest changes are more prevalent, such as in the forest edges in DRC, southern Cameroon, and coastal west Africa (Figure 3, Supplemental Figure 3). The prediction location that had the highest contrast between the full and reduced models in the Zaire-only analysis occurred in DRC where the estimated spillover ROR from the full model was 67.1 times that produced by the reduced model, and 7 countries had at least one 1x1km grid cell whose estimated spillover ROR was >20 times that from the reduced model, including DRC, Cameroon, Liberia, Guinea, Sierra Leone, Cote d’Ivoire, and Ghana (Figure 3, Supplemental Table 4). Incorporation of forest change data into the All-species analysis produced similar results to those from the Zaire-only analysis, however, the magnitude of difference between the full and reduced models was smaller in most countries in the study area, where the prediction grid cell with the highest contrast between the full and reduced models occurred in Cameroon, where estimates were 16.1 times as high in the full model compared to the reduced model, followed by DRC (ratio: 14.9), and Liberia (ratio: 8.2) (Figure 3, Supplemental Table 4).

While including forest change data in the full models led to identification of areas with elevated odds of spillover that were missed in the reduced models, such as in coastal West Africa, it also led to a 59.5% reduction in the total area with estimated spillover RORs >20 in the All-species analysis and a 26.0% reduction in the total area with estimated spillover RORs >20 in the Zaire-only analysis. Adding forest change variables to the model resulted in a change in the direction of relative spillover odds for 10.2% of the study area in the All-species analysis, where 5.5% (n=5334) of prediction locations that had below average spillover odds in the reduced model had above average odds after accounting for forest loss and fragmentation, and 4.8% (n=4631) of locations that had above average spillover odds in the reduced model change to having spillover ROR <1 after accounting for forest loss and fragmentation. Among the 1x1 kilometer prediction locations whose estimated spillover ROR changed from below-to above-average by adding forest change information to the All-species analysis, 87.7% (n=4676) had a spillover ROR from 1-3, 12.0% (n=642) had a spillover ROR from 3-10, and 0.3% (n=16) had a spillover ROR from 10-20. In the Zaire-only analysis, 11.3% of prediction locations had a change the direction of the spillover ROR, where 6.3% (n=6177) of prediction locations changed from having below-average spillover odds in the reduced model to having above-average spillover odds in the full model. In contrast, 5.0% (n=4894) of prediction locations that had above-average spillover odds in the reduced model changed to having below-average spillover odds in the full model. Among prediction locations in the Zaire-only analysis whose estimated spillover ROR changed from <1 in the reduced model to >1 in the full model, 57.9% (n=3575) had a spillover ROR from 1-3, 35.7% (n=2208) had a spillover ROR from 3-10, 5.9% (n=366) had a spillover ROR from 10-20, and 0.5% (n=28) had a spillover ROR estimate >20 (Supplemental Table 5).

We also conducted a sensitivity analysis to identify differences in relative spillover odds estimates when defining forests as locations with >80% forest cover when classifying fragmented forests compared to 70%, which was used for the primary analysis. Spatial trends in estimated spillover RORs were similar across the study area between the two cutoffs to classify fragmented areas. However, a ratio comparison between ROR estimates resulting from both cutoffs showed that when classifying fragmented forests based on a forest definition of 80%, ROR estimates were as much as 5 times as high in western DRC and in southern Cameroon. In contrast, ROR estimates resulting from forest fragmentation based on a forest definition of 80% resulted in lower ROR estimates in northern DRC along forests edges near its borders with South Sudan and Central African Republic, compared to a forest definition of 70% (Supplemental Figure 4).

### Temporal Shifts in Relative Spillover Odds from 2021-2022

In addition to estimating the spillover RORs for the All-species and the Zaire-only analyses for the year 2022, each model was also fitted to covariate data for the year 2021 to identify temporal shifts in the spatial distribution of estimated spillover odds between the two years. Increases in spillover odds estimations from 2021-2022 occurred in locations scattered throughout the study area, and the spatial pattern was similar between both the All-species and Zaire-only analyses, although the magnitude of change in spillover odds was more pronounced for the model of Zaire virus. Regions that saw the largest areas of increase in estimated spillover RORs included southwestern Ghana, southern Sierra Leone, southern Cameroon, southwestern and northeastern DRC, southern Nigeria, Central African Republic, and patches near the western and eastern borders of South Sudan. A notable difference in the estimated shifts in spillover odds between the All-species and Zaire-only analyses occurred in northern Angola and its border with DRC, where the All-species analysis estimated an increase in spillover odds from 2021-2022 and the Zaire-only analysis estimated a decrease in spillover odds over this period. In contrast, estimated spillover odds increased in eastern Liberia according to the Zaire-only analysis, but decreased in the All-species analysis (Figure 4). In the All-species analysis, the location with the largest increase in spillover odds from 2021-2022 occurred in DRC, where the estimated odds of spillover increased by 4.2 times. In the Zaire-only analysis, the location that had the largest increase in spillover odds from 2021 to 2022 occurred in Ghana, where the estimated spillover ROR increased by 5.9 times.

**Figure 4.**
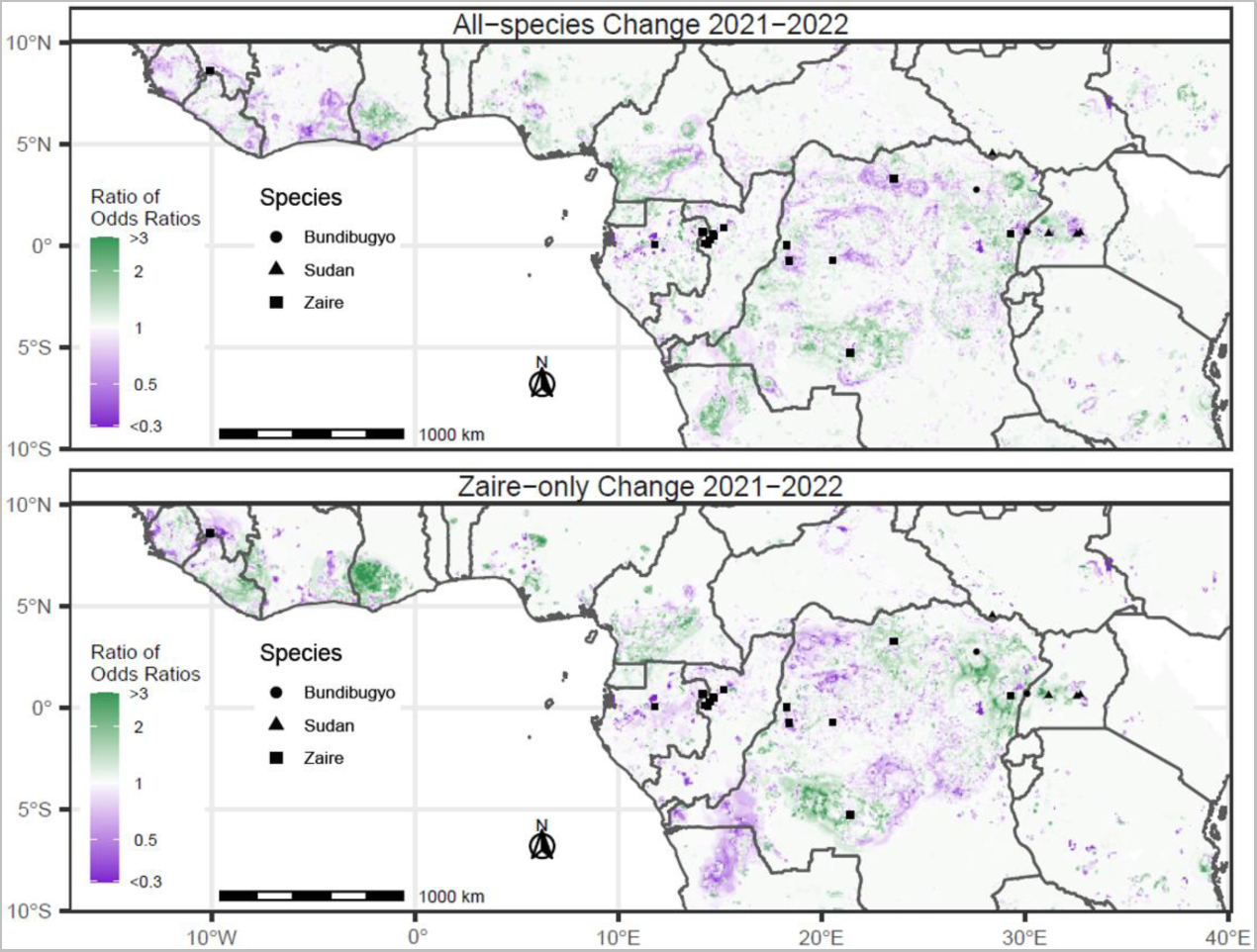
Change in predicted relative odds of spillover from 2021-2022. Ratio between estimated relative spillover odds in 2022 compared to 2021 where green shades represent locations where estimated odds of spillover increased, and purple shades represent locations where estimated relative spillover odds decreased. The top row represents predictions resulting from a model trained on all ebolavirus species combined, while the bottom row represents predictions resulting from a model trained solely on spillover events of Zaire ebolavirus.

### Estimated Spillover Odds at 2022 Spillover Sites

Two zoonotic ebolavirus spillover events were identified in 2022. First, a zoonotic spillover of Zaire ebolavirus was reported in the Mbandaka District of Equateur Province in western DRC. Within Mbandaka District in 2022, the maximum estimated spillover ROR resulting from the Zaire-only analysis was 21.5, which ranked within the top 0.1% in equatorial Africa and the top 0.3% in DRC. Mbandaka District was also the location of the second highest overall 1x1 kilometer prediction cell in equatorial Africa in 2021 (ROR: 31.3). The second zoonotic spillover in 2022 resulted from Sudan ebolavirus in the Mubende District of Uganda, which had a maximum estimated spillover ROR of 2.0, which ranked in the top 12% of spillover RORs resulting from the All-species analysis in 2022 in equatorial Africa, and the top 6% in Uganda. From 2021-2022, a large proportion of Mubende District also saw an increase in estimated spillover RORs; increasing by as much as 2.1 times from 2021-2022, which ranked in the top 0.3% in both equatorial Africa and Uganda (Supplemental Figure 5).

## Discussion

Using data on annual forest cover, forest loss and fragmentation, human population distribution, and climate, we estimate the relative odds of ebolavirus spillover in equatorial Africa in 2022, identify locations where spillover odds estimates were driven by forest loss and fragmentation, highlight temporal shifts in estimated relative spillover odds between 2021-2022, and evaluate estimated spillover odds at locations that actually saw a spillover event in 2022. Using a model trained on spillover events of all ebolavirus species (All-species analysis), the locations with the highest predicted spillover odds included patches throughout central and northern DRC, Gabon, ROC, Cameroon, Nigeria, Central African Republic, Uganda, Ethiopia, Kenya, and coastal west Africa. Predictions resulting from a model trained only on spillover events of Zaire ebolavirus (Zaire-only analysis) followed a similar spatial trend to those from the All-species model, but values of predicted relative odds were higher around DRC, the border between Gabon and ROC, and in coastal west Africa, and lower in areas with less forest cover such as in Uganda, Ethiopia, and Kenya. Comparing spillover odds estimates between full models and models that ignored forest loss and fragmentation data, we found that incorporation of forest change data led to higher estimates of spillover odds along the northern and southern edges of the DRC rainforest, in southern Cameroon, and in several countries in coastal west Africa. Shifts in estimated odds of spillover occurred in patches throughout equatorial Africa and were of greater magnitude in the Zaire-only analysis, with the largest areas of increase occurring in southwestern and northeastern DRC, southern Cameroon, and coastal West Africa.

Past estimates of the spatial distribution of ebolavirus spillover risk have not accounted for the impact that forest loss and fragmentation have on spillover likelihood across varying spatial scales, and most do not consider temporally changing risk given annual shifts in human populations and forests. Publicly available data on forest loss and human population distributions are accessible on an annual basis and fitted parameters from these models can be used to annually update spillover odds estimates and guide public health prevention and risk mitigation activities, target surveillance efforts, and strengthen relationships with local health clinics in high-risk areas to ensure rapid reporting and testing of suspect ebolavirus cases. This study was able to identify specific regions within equatorial Africa with elevated odds of spillover, with approximately 20% of the area having relative spillover odds >1 (11,13–16). While still infeasible to prioritize surveillance and prevention efforts in all regions with above-average odds of ebolavirus spillover, prioritization of locations with the highest estimates of relative spillover odds and those with recent increases in estimates may be beneficial for rapid outbreak detection. For example, spillover events that occurred in 2022 in DRC and Uganda occurred in locations that ranked among those with the highest overall estimated spillover RORs and/or 1-year increases from 2021-2022. Mbandaka District was also the location of the second highest estimate of spillover odds in 2021 (Supplemental Figure 5). While impossible to predict ebolavirus spillover, prioritization of regions with the highest overall odds of spillover and the largest annual increases in estimates would have prioritized the two locations that actually saw a spillover event and could have potentially enabled earlier detection and initiation of public health response. However, prioritization of resource allocation in regions with potentially high-risk should still consider local context that may not agree with our model estimates. For example, the Zaire-only analysis identifies the eastern half of Central African Republic as a region with moderate odds of spillover. Comparison with the covariate distribution maps suggest that this area was identified by the model given that it is a large rural area, and rurality across large spatial scales was among the top predictors of Zaire ebolavirus spillover. However, this is also an area with low forest cover, suggesting that its extreme rurality may have caused excessive inflation of its spillover odds estimates.

Aside from the epidemiological relevance of these findings, ongoing ecological efforts to identify a natural reservoir for ebolaviruses may also benefit by targeted sampling of suspect bat reservoirs at specific times and locations with elevated spillover odds. In areas where human population and animal habitat perturbation will be known to occur, such as where roads or communities will be expanding, studies could evaluate changes in animal behavior and seroprevalence among suspect reservoirs and human populations, especially if environmental changes correlate in time with bat birthing patterns, which have been shown to be correlated with increased filovirus transmission among bat populations (16,40). In comparison to spillover predictors such as vegetation density and temperature, forest loss is a risk driver that could immediately be addressed through implementation and enforcement of environmental policy and provides an actionable target that could potentially reduce risk of future outbreaks of ebolaviruses and many other infectious diseases (41). Potential hypotheses regarding animal reservoir species of each individual ebolavirus species could be generated given the difference in estimated spillover odds between the All-species and Zaire-only analyses. Forest loss variables tended to have higher predictive importance in the Zaire-only analysis, especially when measuring forest loss 1- and 2-years prior and within close proximity (10 kilometers) to spillover sites. Differences in the top predictors of spillover between the two analyses are evident in the contrast between estimated temporal shifts in spillover odds between the two analyses. For example, according to the Zaire-only analysis, spillover odds increased in eastern Liberia and decreased in northern Angola in 2022; whereas according to the All-species analysis, spillover odds decreased in eastern Liberia and increased in northern Angola. Given that the only difference between the All-species and Zaire-only analyses was the inclusion of Sudan and Bundibugyo virus species, it can be hypothesized that unique environmental conditions represented in this model may be associated with emergence of each species of ebolavirus. Under the assumption that each ebolavirus species has a unique animal reservoir species, the natural reservoir of Zaire ebolavirus could potentially be an animal species that is localized to dense forests, is more impacted by disruption of forest habitats, or potentially remains within close proximity to deforested locations rather than immigrate away to unaffected habitats. Little research has been done to evaluate the type of land conversion that takes place in recently deforested areas surrounding ebolavirus spillover sites, and future research should seek to determine the specific land types that replace deforested areas. For example, deforested areas replaced with fruit crops could draw increasing numbers of fruit bats into areas nearby human populations, resulting in elevated likelihood of spillover (42).

According to cross validation AUC values, inclusion of forest loss and fragmentation covariates across scales (full models) resulted in small changes to predictive accuracy in both analyses. Though the prediction accuracy in the Zaire-only analysis was only marginally higher in the full model, this improvement suggests that forest loss and fragmentation variables may represent underlying environmental dynamics associated with ebolavirus spillover, especially for Zaire ebolavirus. In contrast, inclusion of Sudan and Bundibugyo viruses with Zaire virus in the All-species analysis resulted in poorer prediction accuracy, suggesting that forest loss and fragmentation may be less important to the spillover ecology of Sudan and/or Bundibugyo viruses. It was not possible to conduct individual Sudan-only and Bundibugyo-only analyses given the low number of their respective emergences from 2001-2021 (Sudan=4, Bundibugyo=2). While inclusion of forest change variables resulted in only small changes to prediction accuracy in both the All-species and Zaire-only analyses, it is notable that when they were included, these variables were identified as some of the predictors with the highest relative importance. As a result, clear differences are evident in the maps of estimated relative spillover odds when using full models compared to reduced models in both analyses. For example, coastal west Africa was estimated to have below-average odds of spillover by the reduced model in the Zaire-only analysis, notwithstanding Guinea being the site of a spillover of Zaire ebolavirus in 2014. However, after accounting for forest change variables in the full model, large proportions of Sierra Leone, Liberia, and the southern border of Guinea were estimated to have spillover RORs >3. These results suggest that inclusion of forest loss and fragmentation provide valuable information that could encompass an important element of the spillover ecology of ebolaviruses, especially Zaire ebolavirus.

We present estimates of the spatial distribution of ebolavirus spillover as the odds of spillover relative to the overall mean odds in the study area, and it is important to note that ebolavirus emergence is a rare outcome and the true odds of spillover are low, even in areas with odds that are high relative to the average odds across the study area.

Therefore, most points classified as having the potential for emergence will typically not actually be places where the disease emerges. For instance, the odds cutoff in the ROC curve for the full model of the All-species analysis that best balanced sensitivity and specificity was 0.0084, which produced a sensitivity of 86% and specificity of 73%. Therefore, any prediction location with spillover odds greater than 0.0084 would have been classified as a potential spillover site, when in reality most of these locations would never actually experience ebolavirus spillover. In the All-species analysis full model, there were 2,750 locations that were classified as potential spillover sites, only 19 of which ever saw an emergence event over the 20-year study period from 2001-2021. Similarly for the Zaire-only analysis, using the cutoff that best balanced sensitivity and specificity (cutoff: 0.0159, sensitivity: 94%, specificity: 87%), there were 1,360 points classified as having high potential for spillover, and only 15 actually saw an emergence event from 2001-2021 (Figure 1). Predictive accuracy also varies depending on the geographic region that is analyzed. Individual country level analyses would likely result in lower predictive accuracy because presence and absence points sampled from within a smaller area would have greater homogeneity in environmental conditions, resulting in reduced predictive sensitivity. Given that all known spillover events of ebolaviruses have occurred in forests near the African equator, we limited our study area and generation of absence locations within 10 degrees latitude of the equator and to locations with an annual average precipitation >500mm, rather than include large proportions of the African continent as has been done previously (14,15).

These analyses have several limitations. First, geographic coordinates for spillover events represented the village of residence of human index cases of ebolavirus outbreaks. Therefore, our predictions represent locations where an index case of an outbreak is likely to be identified and not necessarily the location where spillover occurred. Our incorporation of various spatial scales of data aggregation is conducive to this limitation, as larger spatial scales surrounding the village of an index case are meant to encompass areas where infection may have occurred. Second, covariates representing forest loss during the same year as the spillover event may include forest loss in months following spillover events, as forest loss data are reported annually, and it could not be determined whether forest loss occurred prior to and after a spillover event in the same year. Third, forest cover data were available beginning in 2000, allowing calculation of forest loss beginning in 2001. Therefore, we were not able to analyze spillover events from 1975-2000 and could not include forest loss temporal lags for spillover events that occurred in 2001 and could only analyze a forest loss temporal lag of 1 year for spillover events that occurred in 2002. Finally, while we found forest fragmentation to be an important predictor of ebolavirus spillover, its relationship with spillover could change based on how “fragmented forests” are defined. We used the method outlined by *Riitters et al.* which requires a binary definition of “forests" prior to determining which areas surrounding forests would be considered “fragmented” (36). We used cutoffs of 70% and 80% to define “forests” prior to classifying fragmented areas and found that while spatial trends remained similar between the two cutoffs, relative spillover odds estimates were higher in the center of the African rainforest when using a cutoff of 80%, and higher along forests edges when using a cutoff of 70% (Supplemental Figure 4). Using lower cutoffs to define “forests” prior to classifying fragmented areas would therefore likely result in an expanded area of fragmented forests further from the center of the equatorial African rainforest and potentially identify additional areas with potential for spillover. The optimal cutoff to define “forests” prior to classifying fragmented areas would likely vary across pathogens and geography based on pathogen ecology and transmission dynamics, and given that ebolavirus spillover has historically occurred in proximity to dense forests, we chose to limit our forest cutoff definitions to relatively high values.

Ebolaviruses continue to be a relevant threat to human and animal health, and continued research is needed to understand the process leading to zoonotic spillover. We found that variables representing forest loss, fragmentation, and human population had relationships with ebolavirus spillover, and inclusion of forest loss and fragmentation considerably changed the geographic distribution of ebolavirus spillover estimates. However, our understanding of the relationship between forest loss, fragmentation, and ebolavirus remains poor, and will likely continue to be limited until a ebolavirus reservoir is confirmed. Results in these analyses could be used to target public health surveillance efforts to rapidly identify and respond to ebolavirus outbreaks and guide ecological research seeking to confirm an ebolavirus reservoir.

## Methods

### Ebolavirus spillover data

Geographic coordinates for known spillover events of ebolavirus into humans from 2001-2022 were ascertained from published literature, reports of outbreaks from the Centers for Disease Control and Prevention (CDC) and World Health Organization, and through consultation with CDC outbreak responders who traveled to the villages of residence of index cases of outbreaks. Spillover events prior to 2001 were excluded because annual forest cover, forest loss, and human population count data were not available prior to this year. We conducted geographic analysis using centroid coordinates for the village of residence of human index cases of outbreaks, assuming infection occurred in proximity to this location. A total of 24 unique ebolavirus spillover events were identified from epidemiologic outbreak investigations and included in this analysis, among which 17 were associated with the Zaire species, 5 with the Sudan species, and 2 with the Bundibugyo species (Supplemental Figure 3). Recent evidence has shown the possibility of transmission among latently infectious survivors of past outbreaks (43). Given that such events are not related to zoonotic transmission from a natural reservoir, we excluded such cases from this analysis.

### Covariate data

All model covariates were ascertained within a study area encompassing ebolavirus spillover events from 2001-2022 within 10 degrees latitude of the equator on the African continent (Supplemental Figure 3). Only locations with a historical average >500mm of annual precipitation were included, as past spillover events have only been identified in areas with high precipitation and vegetation density (15). Forest cover data were available from Hansen et al. at a spatial resolution of 30x30m for the year 2000 and subsequent annual forest loss from 2001-2022 (35). Individual maps of annual forest cover from 2000-2022 were generated and used to calculate annual percent forest loss at a spatial resolution of 1x1km. Annual forest loss prior to a spillover event was analyzed as the difference in forest cover percentage across three temporal scales: the same year, one year prior, and two years prior to each spillover event. Past research has found that fragmented forest such as forest edges and patches are associated with ebolavirus spillover (17). We analyzed by classifying forest fragmentation according to methods outlined by *Riitters et al.*, considering patch, transitional, edge, and perforated areas to be fragmented (36). Because of the potential for forest fragmentation results to vary by the cutoff used to define forests, we tested cutoffs of 70% and 80% to classify forest fragmentation (17,36). Annual human population distribution data were ascertained from Landscan and log transformed due to the skewed distribution of human populations (44). An interaction variable combining log human population count and forest cover was created by multiplying the two values to represent locations with both high numbers of susceptible individuals and high vegetation density. Other covariates were analyzed as historical annual averages, which included potential evapotranspiration (PET), night-time land surface temperature (NTLST), elevation, temperature seasonality (standard deviation of monthly temperature), and precipitation seasonality (coefficient of variation of monthly precipitation). A full list of covariates along with their corresponding sources, spatial resolution, collection frequency, and units of measurement is available in Supplemental Table 2.

### Statistical analysis

For comparison with spillover events in models, we randomly generated 10,000 pseudo-absence points (absence locations) within the study area and randomly assigned a year from 2001-2021 to each absence location, thereby representing a time and location at which an ebolavirus spillover was *not* identified. Absence locations were generated with replacement and biased toward areas with increased log population count to account for increased reporting in populated areas. Mean values for percentage in forest loss (same year, one year prior, two years prior), forest cover, and fragmentation were extracted at each presence and absence location across five spatial scales using mutually exclusive donut buffers with maximum radii of 10, 25, 50, 100, and 150 kilometers. Human population count was extracted as the log transformed sum of humans within each of the five spatial scales. Elevation, PET, NTLST, temperature seasonality, and precipitation seasonality were not analyzed across scales but rather extracted as the mean within 10 kilometers of each presence and absence location. The interaction between log human population count and forest cover was calculated using the data extracted within the 10 kilometers spatial scale. Therefore, the model included 36 covariates (30 scaled + 6 non-scaled). A descriptive analysis compared mean and standard deviation of covariate values between spillover and absence locations.

Models estimating the odds of ebolavirus spillover were fit using boosted regression trees (BRT) with a Bernoulli error distribution for binomial outcomes using the seegSDM package using R software (45,46). BRT quantifies the relationship between the outcome and predictor variables by generating regression trees with binary splits of randomly sampled predictor values. Regression trees are iteratively generated based on residuals from the previous tree, resulting in the gradual reduction in prediction residuals as subsequent trees are built. BRT then combines the information from each tree to quantify the relationships between the outcome and each covariate and make predictions (47). Trees based machine learning algorithms are robust methods for analysis of many covariates as they analyze combinations of subsets of covariates, thus reducing bias when correlation exists between covariates, as could be the case for our scaled covariates. BRT also allows for analysis of covariates with missing data values by generating a split for missing data in the regression trees, which accommodated missing historical forest loss data when analyzing one- and two-year lag periods between forest loss and spillover events occurring from 2001-2002.

Two separate predictive analyses were run based on spillover events occurring from 2001-2021; one which included spillover events of all ebolavirus species (All-species analysis), and a second which included only spillover of Zaire ebolavirus (Zaire-only analysis). For each analysis, an ensemble of 100 models was used, each of which randomly sampled 50 pseudo-absence locations (from the possible 10,000) for each presence location with replacement. Hyperparameters for both models were selected using a tuning grid to test optimal tree depth, learning rate, and bag fraction. We calculated area under the receiver operator curve (AUC) using 3-fold cross validation to evaluate the predictive ability of each model ensemble. Folds were assigned to spillover sites in descending order through time, such that each fold contained spillover sites distributed throughout the study period, and absence points were randomly assigned to one of the 3 folds. For each fold, the odds of spillover at each spillover and absence location were estimated based on a model ensemble fit to the remaining two folds. Fitted values from the models were output on the odds scale. Plots were generated to visualize the relative predictive importance of each covariate and the marginal response curves for the 12 most important predictors for each analysis. Models were then fit to covariate values on a 10x10 kilometer prediction grid of the study area for the years 2021 and 2022. For each prediction location, we calculated the relative odds of spillover as the fitted odds of spillover divided by the mean odds of spillover across the entire study area. To quantify the contribution of forest loss and fragmentation to estimated relative spillover odds, a reduced version of each model was run again excluding forest loss and fragmentation covariates to visualize the impact of these variables on estimated spillover odds across the study area. Models reported in the main body of this manuscript use forest fragmentation measures using 70% as the cutoff definition for forest cover, and model results using 80% forest cover as the cutoff to classify fragmented forest are reported in the supplementary materials.

### Competing interests

The authors have declared that no competing interests exist.

The opinions expressed by the authors do not necessarily reflect the opinions of the Centers for Disease Control and Prevention or the institutions with which the authors are affiliated.

## Data Availability

All data produced in the present study are available upon reasonable request to the authors

---

**Supplemental Table 1.** **Ebolavirus spillover event metadata.**

| *ID* | *Country* | *Ebolavirus Species* | *Year* | *Lat* | *Long* | *Source* |
| --- | --- | --- | --- | --- | --- | --- |
| 1 | Gabon | Zaire | 2001 | 0.0550 | 11.8109 | Lahm et al 2007, Nkoghe et al. 2011 |
| 2 | Gabon | Zaire | 2001 | 0.7001 | 14.1580 | Leroy et al. 2002, Allela et al. 2005, Pourrut et al. 2005 |
| 3 | ROC | Zaire | 2002 | 0.1333 | 14.2667 | Pourrut 2005, Nkoghe et al. 2011 |
| 4 | ROC | Zaire | 2002 | 0.2987 | 14.5075 | Rouquet et al. 2005 |
| 5 | ROC | Zaire | 2003 | 0.0682 | 14.4200 | Pourrut 2005, Nkoghe et al. 2011, Mylne et al. 2014 |
| 6 | ROC | Zaire | 2003 | 0.5619 | 14.6573 | Pourrut 2005 |
| 7 | South Sudan | Sudan | 2004 | 4.5568 | 28.4016 | https://apps.who.int/iris/bitstream/handle/10665/232925/WER8043_370-375.PDF |
| 8 | ROC | Zaire | 2004 | 0.9064 | 15.1751 | Caillaud et al. 2006 |
| 9 | ROC | Zaire | 2005 | 0.4944 | 14.6786 | CDC responder retrospective identification of case village |
| 10 | DRC | Zaire | 2007 | -5.2596 | 21.4095 | Nkoghe et al. 2011 |
| 11 | Uganda | Bundibugyo | 2007 | 0.7038 | 30.1175 | Leroy et al. 2009, Mylne et al. 2014, Pigott et al. 2014 |
| 12 | DRC | Zaire | 2008 | -5.2401 | 21.4103 | Shoemaker et al. 2012, Mylne et al. 2014 |
| 13 | Uganda | Sudan | 2011 | 0.6444 | 32.7276 | Grard et al. 2011, Muyembe-Tamfum et al. 2012, Mylne et al. 2014 |
| 14 | Uganda | Sudan | 2012 | 0.6214 | 31.1685 | Shoemaker et al. 2012, Mylne et al. 2014 |
| 15 | DRC | Bundibugyo | 2012 | 2.7718 | 27.6196 | Mylne et al. 2014 |
| 16 | Uganda | Sudan | 2012 | 0.5784 | 32.5480 | Epelboin et al. 2012, Pigott et al. 2014 |
| 17 | Guinea | Zaire | 2013 | 8.6225 | -10.0642 | Albarino et al. 2013, Mylne et al. 2014 |
| 18 | DRC | Zaire | 2014 | -0.7139 | 20.5302 | Baize et al. 2014 |
| 19 | DRC | Zaire | 2017 | 3.2990 | 23.5430 | Maganga et al. 2014, Mylne et al. 2014 |
| 20 | DRC | Zaire | 2018 | -0.7368 | 18.4214 | Perma.cc Record, https://perma.cc/PG7D-PEGQ |
| 21 | DRC | Zaire | 2018 | 0.6059 | 29.3065 | CDC responder retrospective identification of case village |
| 22 | DRC | Zaire | 2020 | 0.0300 | 18.2800 | CDC responder retrospective identification of case village |
| 23 | DRC | Zaire | 2022 | 0.0414 | 18.2770 | CDC responder retrospective identification of case village |
| 24 | Uganda | Sudan | 2022 | 0.6697 | 31.4686 | CDC responder retrospective identification of case village |

**Supplemental Table 2.** **Model Covariates**

| Variable | Units | Original Spatial Resolution | Years Represented in Data | Temporal Range of analysis | Citation |
| --- | --- | --- | --- | --- | --- |
| Elevation | Meters | 1x1km | 2000 | Single time point | Fick, S.E. and R.J. Hijmans, 2017. WorldClim 2: new 1km spatial resolution climate surfaces for global land areas. International Journal of Climatology 37 (12): 4302-4315. |
| Forest Cover (FC) | Percentage | 30x30m | 2000-2022 | % FC during year of spillover | Hansen, M. C., Potapov, P. V., Moore, R., Hancher, M., Turubanova, S. A., Tyukavina, A., ... & Townshend, J. (2013). High-resolution global maps of 21st-century forest cover change. science, 342(6160), 850-853 |
| Forest Fragmentation | Binary | 30x30 (Calculated using FC data) | 2000-2022 | % Fragmentation during year of spillover | Riitters K, Wickham J, O'Neill R, Jones B, Smith E. Global-scale patterns of forest fragmentation. Conserv Ecol. 2000;4(2). |
| Forest loss | Binary | 30x30m | 2001-2022 | % forest loss during same year, 1 year prior, and 2 years prior to spillover | Hansen, M. C., Potapov, P. V., Moore, R., Hancher, M., Turubanova, S. A., Tyukavina, A., ... & Townshend, J. (2013). High-resolution global maps of 21st-century forest cover change. science, 342(6160), 850-853. |
| Night-time Land Surface Temperature (NTLST) | Degrees Kelvin | 0.05 degrees | 2000-2022 | Time Averaged mean monthly values | Wan, Z., Hook, S., Hulley, G. (2015). MOD11C3 MODIS/Terra Land Surface Temperature/Emissivity Monthly L3 Global 0.05Deg CMG V006 [Data set]. NASA EOSDIS Land Processes DAAC. Accessed 2021-10-11 from https://doi.org/10.5067/MODIS/MOD11C3.006 |
| Potential Evapotranspiration (PET) | mm day-1 | 1x1km | 2000-2022 | Time Averaged mean monthly values | Trabucco, A., and Zomer, R.J. 2018. Global Aridity Index and Potential Evapo-Transpiration (ET0) Climate Database v2. CGIAR Consortium for Spatial Information (CGIAR-CSI). Published online, available from the CGIAR-CSI GeoPortal at https://cgiarcsi.community |
| Precipitation | Millimeters | 1x1km | 1970-2000 | Time Averaged mean monthly values | Fick, S.E. and R.J. Hijmans, 2017. WorldClim 2: new 1km spatial resolution climate surfaces for global land areas. International Journal of Climatology 37 (12): 4302-4315. |
| Temperature Seasonality | Standard Deviation | 1x1km | 1970-2000 | Time Averaged mean monthly values | Fick, S.E. and R.J. Hijmans, 2017. WorldClim 2: new 1km spatial resolution climate surfaces for global land areas. International Journal of Climatology 37 (12): 4302-4315. |
| Precipitation Seasonality | Coefficient of Variation | 1x1km | 1970-2000 | Time Averaged mean monthly values | Fick, S.E. and R.J. Hijmans, 2017. WorldClim 2: new 1km spatial resolution climate surfaces for global land areas. International Journal of Climatology 37 (12): 4302-4315. |
| Population count | Population per grid cell | 30 arc seconds (~1xkm) | 2000-2022 | Time Averaged mean annual values | LandScan (2000-2022)<sup>TM</sup> High Resolution Global Population Data Set copyrighted by UT-Battelle, LLC, operator of Oak Ridge National Laboratory under Contract No. DE-AC05-00OR22725 with the United States Department of Energy. |

**Supplemental Table 3.**
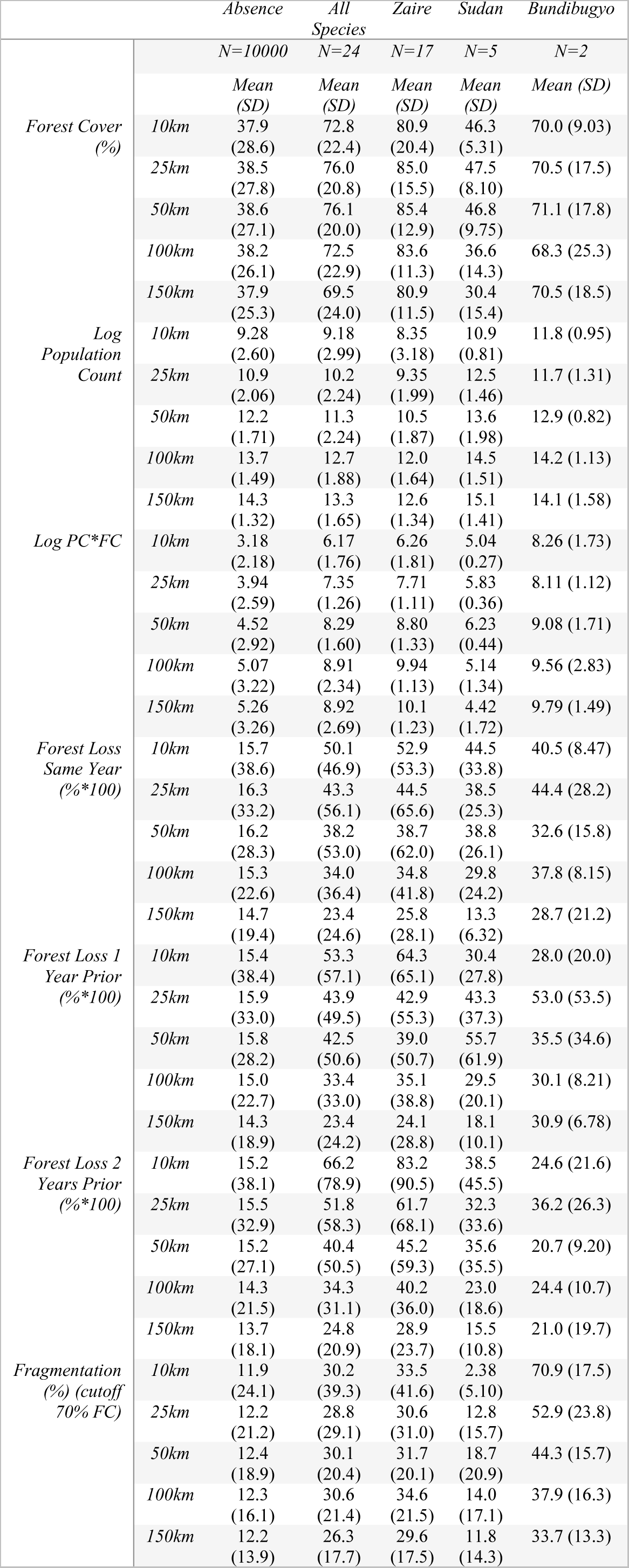
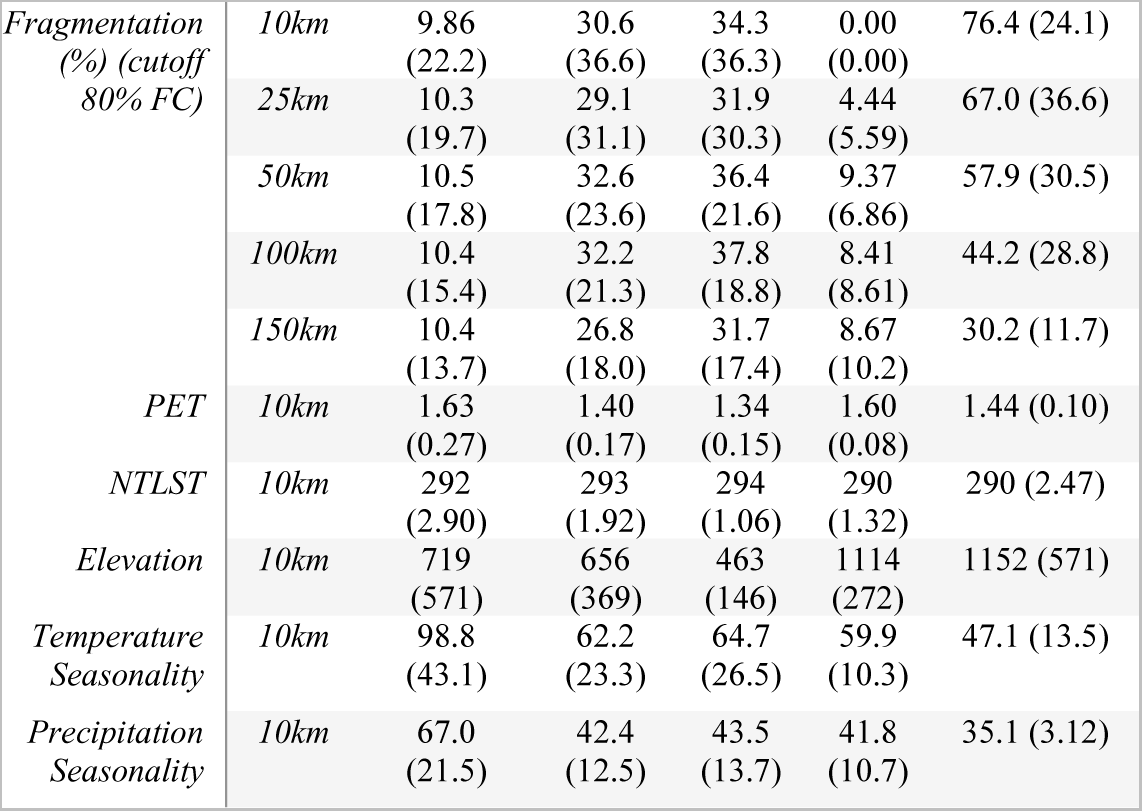
**Descriptive analysis of covariates stratified by ebolavirus species.**

**Supplemental Table 4.** **Comparison of relative spillover odds estimates between the All-species and Zaire-only analyses.**

|  | All-species Analysis |  |  |  | Zaire-only Analysis |  |  |  | Maximum Ratios |  |  |  |  |  |
| --- | --- | --- | --- | --- | --- | --- | --- | --- | --- | --- | --- | --- | --- | --- |
|  | Full Model |  | Reduced Model |  | Full Model |  | Reduced Model |  | All-species Analysis |  |  | Zaire-only Analysis |  |  |
| Country | Mean Spillover ROR | Max. spillover ROR | Mean Spillover ROR | Max. spillover ROR | Mean Spillover ROR | Max. spillover ROR | Mean Spillover ROR | Max. spillover ROR | All-species / Zaire-only ROR estimates (Full models) | Full / Reduced model ROR Estimates | 2022 / 2021 ROR Estimates (Full models) | Zaire-only / All-species ROR estimates (Full models) | Full / Reduced model ROR Estimates | 2022 / 2021 ROR Estimates (Full models) |
| Angola | 0.50 | 5.27 | 0.41 | 4.94 | 0.29 | 3.51 | 0.27 | 2.07 | 15.79 | 7.54 | 4.17 | 1.49 | 12.79 | 2.29 |
| Benin | 0.32 | 1.01 | 0.35 | 0.77 | 0.17 | 0.66 | 0.23 | 0.28 | 5.01 | 1.51 | 2.31 | 1.50 | 2.91 | 2.25 |
| Burkina Faso | 0.29 | 0.29 | 0.34 | 0.34 | 0.17 | 0.17 | 0.23 | 0.23 | 1.72 | 0.87 | 1.03 | 0.58 | 0.75 | 1.02 |
| Burundi | 0.46 | 3.37 | 0.71 | 9.16 | 0.17 | 0.29 | 0.28 | 1.40 | 16.90 | 1.05 | 1.81 | 0.62 | 0.77 | 1.04 |
| Cameroon | 1.12 | 20.83 | 0.76 | 35.83 | 0.94 | 23.40 | 0.54 | 25.72 | 13.93 | 16.70 | 3.98 | 2.38 | 44.50 | 3.01 |
| Central African Rep. | 1.03 | 10.21 | 0.98 | 14.19 | 1.45 | 10.27 | 1.37 | 9.17 | 9.47 | 7.55 | 2.32 | 2.63 | 16.44 | 2.67 |
| Chad | 0.32 | 2.35 | 0.36 | 2.54 | 0.21 | 3.94 | 0.27 | 3.99 | 2.17 | 2.42 | 1.64 | 1.83 | 5.35 | 1.89 |
| Congo | 1.12 | 28.14 | 1.88 | 44.03 | 1.14 | 23.56 | 2.20 | 39.86 | 6.44 | 7.89 | 3.00 | 3.88 | 10.39 | 3.28 |
| Cote d'Ivoire | 0.45 | 4.37 | 0.36 | 3.09 | 0.33 | 6.35 | 0.23 | 0.52 | 7.27 | 7.01 | 1.91 | 3.21 | 25.73 | 3.08 |
| Dem. Rep. of Congo | 1.96 | 32.33 | 1.72 | 42.37 | 2.09 | 31.18 | 1.84 | 50.68 | 24.87 | 14.86 | 4.18 | 5.09 | 67.14 | 5.40 |
| Equatorial Guinea | 0.78 | 12.54 | 1.95 | 11.18 | 0.40 | 7.43 | 2.03 | 8.44 | 6.41 | 2.65 | 2.82 | 1.44 | 2.33 | 2.24 |
| Ethiopia | 0.40 | 3.40 | 0.65 | 12.27 | 0.18 | 1.21 | 0.28 | 1.97 | 10.01 | 1.97 | 2.74 | 1.36 | 2.93 | 1.65 |
| Gabon | 1.67 | 25.94 | 2.84 | 48.63 | 1.84 | 23.56 | 4.09 | 51.76 | 5.89 | 3.94 | 3.50 | 3.88 | 5.25 | 2.99 |
| Ghana | 0.57 | 7.61 | 0.41 | 3.37 | 0.48 | 5.29 | 0.24 | 0.63 | 8.84 | 6.39 | 2.63 | 3.06 | 23.08 | 5.90 |
| Guinea | 0.68 | 7.80 | 0.36 | 3.53 | 0.72 | 6.56 | 0.23 | 0.72 | 4.10 | 7.65 | 1.59 | 2.65 | 29.12 | 2.90 |
| Kenya | 0.37 | 5.78 | 0.56 | 24.98 | 0.18 | 0.43 | 0.25 | 1.78 | 25.33 | 1.72 | 1.85 | 0.73 | 1.55 | 1.19 |
| Liberia | 1.68 | 6.16 | 0.69 | 5.02 | 2.59 | 7.60 | 0.31 | 1.58 | 9.18 | 8.23 | 1.74 | 3.21 | 33.76 | 3.26 |
| Malawi | 0.30 | 0.36 | 0.34 | 0.36 | 0.17 | 0.18 | 0.23 | 0.28 | 2.06 | 1.07 | 1.05 | 0.61 | 0.78 | 1.05 |
| Nigeria | 0.42 | 5.55 | 0.49 | 8.03 | 0.20 | 1.76 | 0.25 | 1.33 | 25.58 | 4.82 | 3.92 | 1.70 | 7.84 | 3.48 |
| Rwanda | 0.57 | 3.78 | 1.38 | 18.85 | 0.26 | 2.73 | 1.09 | 7.85 | 16.83 | 1.32 | 1.85 | 1.01 | 1.68 | 1.08 |
| Sierra Leone | 1.23 | 3.89 | 0.35 | 2.08 | 2.07 | 6.41 | 0.23 | 0.95 | 4.10 | 7.56 | 1.73 | 3.14 | 28.45 | 2.05 |
| South Sudan | 0.40 | 2.33 | 0.45 | 4.77 | 0.35 | 4.33 | 0.42 | 4.05 | 7.46 | 2.86 | 2.44 | 2.27 | 6.07 | 4.59 |
| Sudan | 0.66 | 2.24 | 0.65 | 1.88 | 0.96 | 4.40 | 0.95 | 3.88 | 1.74 | 1.27 | 1.09 | 2.43 | 1.16 | 1.20 |
| Togo | 0.30 | 0.59 | 0.34 | 1.13 | 0.17 | 0.54 | 0.23 | 0.28 | 3.45 | 1.51 | 1.70 | 1.19 | 2.38 | 1.66 |
| Uganda | 0.53 | 13.18 | 0.93 | 35.96 | 0.21 | 1.77 | 0.32 | 8.92 | 53.23 | 3.20 | 2.96 | 1.56 | 7.88 | 3.06 |
| Tanzania | 0.33 | 3.05 | 0.37 | 4.59 | 0.20 | 2.44 | 0.26 | 2.02 | 7.04 | 6.60 | 2.76 | 1.58 | 10.81 | 3.17 |
| Zambia | 0.32 | 0.83 | 0.34 | 0.36 | 0.18 | 0.68 | 0.23 | 0.27 | 2.51 | 2.44 | 1.86 | 1.15 | 3.03 | 1.81 |

**Supplemental Table 5.** **Number and proportion of prediction locations across levels of relative spillover odds ratios, All-species and Zaire-only analyses.**

| Estimated relative Odds Ratio | All-species Analysis Prediction Locations (2022) |  | Zaire-only Analysis Prediction Locations (2022) |  |
| --- | --- | --- | --- | --- |
|  | Full model (%) | Reduced model (%) | Full model (%) | Reduced model (%) |
| <1 | 76328 (78.4) | 77031 (79.2) | 78254 (80.4) | 79537 (81.7) |
| 1 to 3 | 14453 (14.9) | 14466 (14.9) | 10165 (10.4) | 9653 (9.9) |
| 3 to 10 | 5538 (5.7) | 4835 (5.0) | 7234 (7.4) | 6494 (6.7) |
| 10 to 20 | 870 (0.9) | 694 (0.7) | 1365 (1.4) | 1285 (1.3) |
| >20 | 111 (0.1) | 274 (0.3) | 245 (0.3) | 331 (0.3) |

**Supplemental Figure 1.**
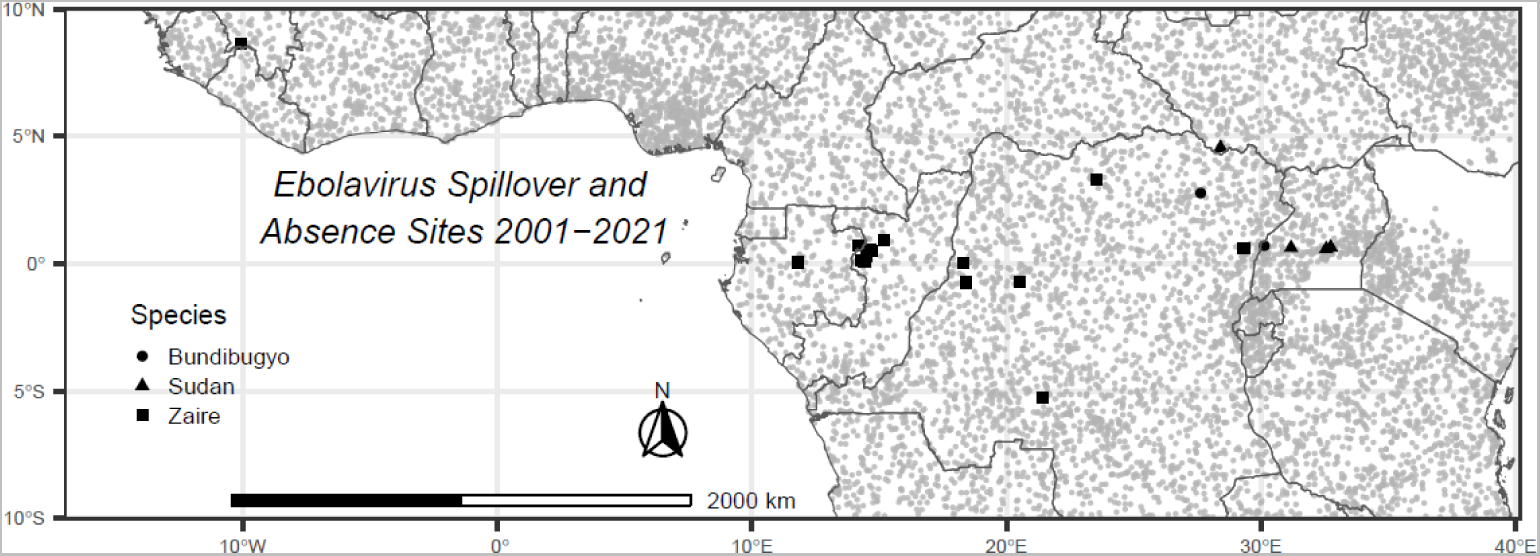
Pseudo absence locations. Gray dots represent pseudo-absence locations randomly generated across the study area and randomly assigned a year from 2001-2021 where ebolavirus spillover was not identified. Pseudo absence generation was biased according to log human population count to account for possible surveillance bias in historical ebolavirus surveillance. Black figures represent ebolavirus spillover sites from 2001-2021

**Supplemental Figure 2.**
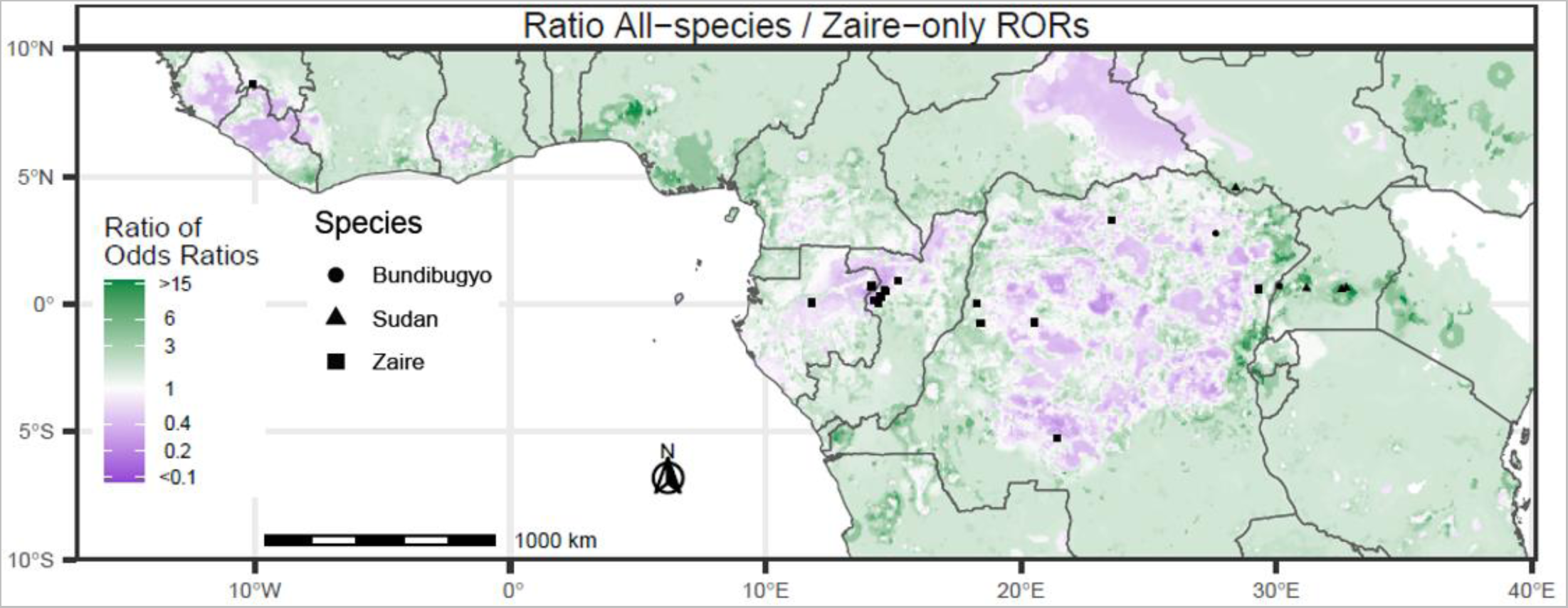
Ratio between full model spillover ROR estimates between All-species and Zaire-only analyses. Areas in green are those that had higher estimated spillover ROR in the All-species analysis and those in purple are those with higher estimated spillover ROR in the Zaire-only analysis.

**Supplemental Figure 3.**
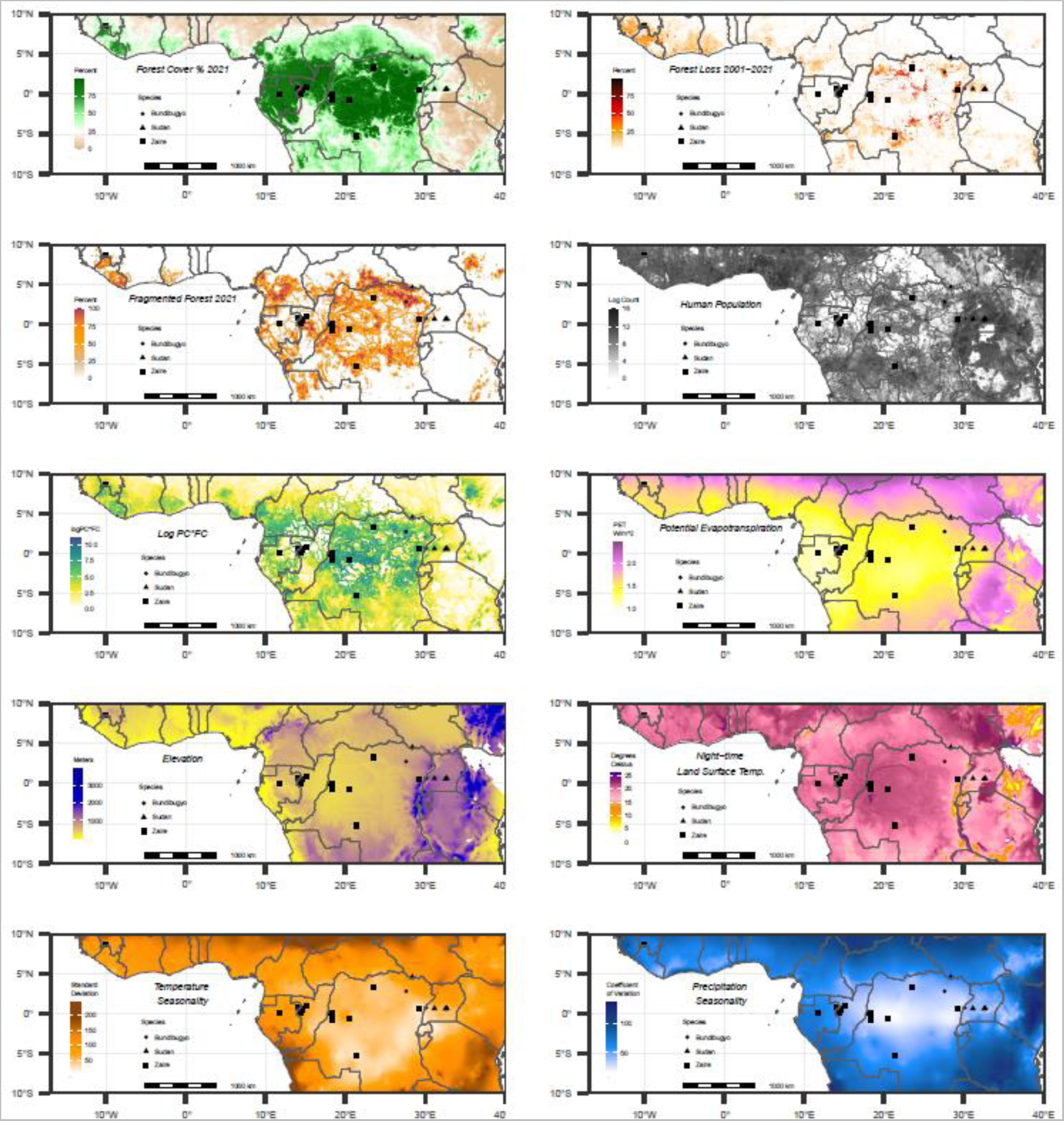
Ebolavirus spillover sites from 2001-2021 and environmental predictors of spillover used to estimate relative odds of spillover. Ebolavirus spillover odds was modeled using forest cover, forest fragmentation, and log human population count during the same year as spillover events, but is visualized here as the values for the year 2021. Annual forest loss was extracted within temporal lags, but is visualized here as total percentage of forest loss from 2001-2021. Meteorological variables including potential evapotranspiration, elevation, night-time land surface temperature, temperature seasonality, and precipitation seasonality were extracted as historical average values across periods outlined in Supplemental Table 2.

**Supplemental Figure 4.**
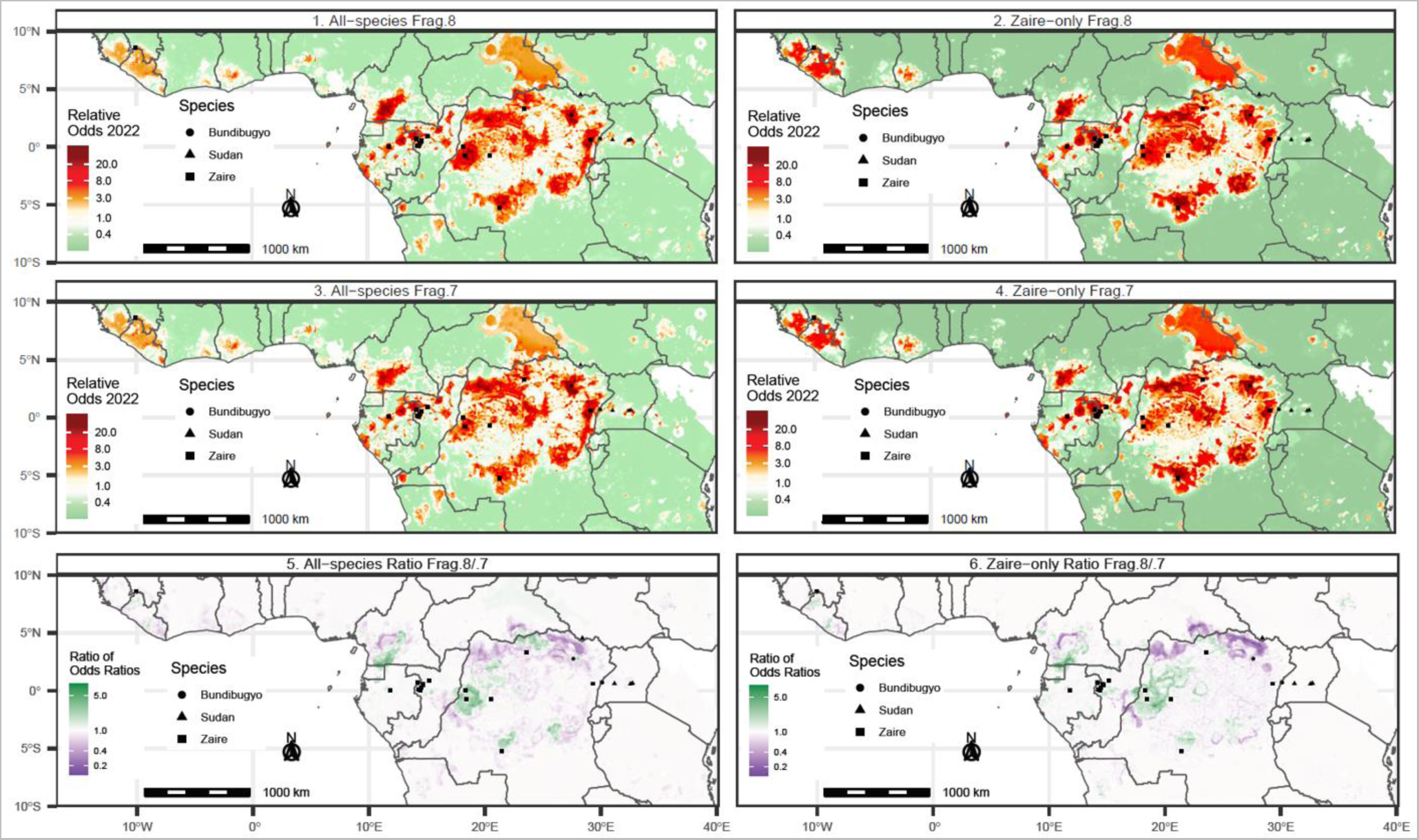
Sensitivity analysis of different forest definition cutoff values used to classify fragmented forests and their impact on relative spillover odds estimates. Based on methods outlined by Riitters et al., fragmented forests are determined based on a binary classification of forests. In the top row are estimates of spillover RORs when defining forests as >80% forest cover when classifying fragmented areas. The middle row represents results presented in the main analysis, where 70% was used to define forests, and subsequently classify fragmented areas. The bottom row represents the ratio between the results from the two different cutoffs, where green represents locations where estimates were higher when forest were defined as >80% forest cover. Purple areas are those that had higher estimates when forests were defined as >70% forest cover. The left column represents results for the All-species analysis, and the right column represents results from the Zaire-only analysis.

**Supplemental Figure 5.**
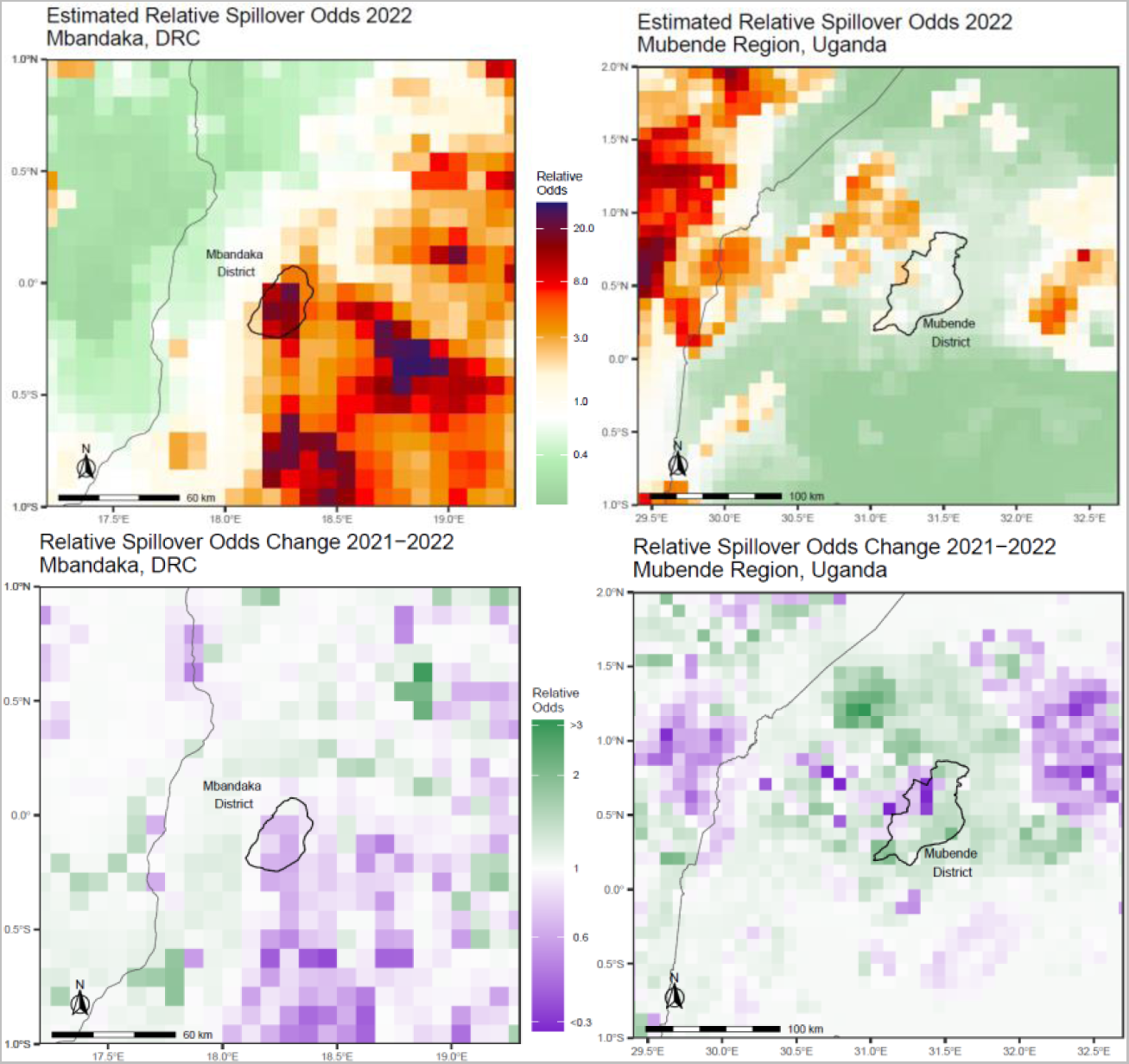
Estimated ebolavirus spillover odds in two locations where ebolavirus spillover events occurred in 2022. Estimated relative odds of ebolavirus spillover produced by the Zaire-only analysis in the Mbandaka health zone in DRC in 2022 (top-left) and the All-species analysis in the Mubende district in Uganda in 2022 (top-right). Changes in the estimated relative odds in each location from 2021 to 2022 are shown on the bottom row, where green shades represent increases in the relative odds of spillover in 2022 relative to 2021 estimates, and purple represents decreases.

## References

1. Rouquet P, Froment JM, Bermejo M, Kilbourn A, Karesh W, Reed P, et al. Wild Animal Mortality Monitoring and Human Ebola Outbreaks, Gabon and Republic of Congo, 2001–2003. Emerg Infect Dis. 2005 Feb;11(2):283–90.

2. Lahm SA, Kombila M, Swanepoel R, Barnes RFW. Morbidity and mortality of wild animals in relation to outbreaks of Ebola haemorrhagic fever in Gabon, 1994–2003. Trans R Soc Trop Med Hyg. 2007 Jan 1;101(1):64–78.

3. Leroy EM, Epelboin A, Mondonge V, Pourrut X, Gonzalez JP, Muyembe-Tamfum JJ, et al. Human Ebola outbreak resulting from direct exposure to fruit bats in Luebo, Democratic Republic of Congo, 2007. Vector-Borne Zoonotic Dis. 2009;9(6):723–8.

4. Nkoghe D, Formenty P, Leroy EM, Nnegue S, Edou SY, Ba JI, et al. Multiple Ebola virus haemorrhagic fever outbreaks in Gabon, from October 2001 to April 2002. Bull Soc Pathol Exot 1990. 2005;98(3):224–9.

5. Plowright RK, Eby P, Hudson PJ, Smith IL, Westcott D, Bryden WL, et al. Ecological dynamics of emerging bat virus spillover. Proc R Soc B Biol Sci. 2015;282(1798):20142124.

6. Nyakarahuka L, Whitmer S, Klena J, Balinandi S, Talundzic E, Tumusiime A, et al. Detection of Sporadic Outbreaks of Rift Valley Fever in Uganda through the National Viral Hemorrhagic Fever Surveillance System, 2017–2020. Am J Trop Med Hyg. 2023;108(5):995.

7. History of Ebola Disease Outbreaks | History | Ebola (Ebola Virus Disease) | CDC [Internet]. 2023 [cited 2023 Aug 29]. Available from: https://www.cdc.gov/vhf/ebola/history/chronology.html

8. History of Ebola Virus Disease (EVD) <B>Outbreaks Error processing SSI file</B><BR> [Internet]. 2021 [cited 2021 Sep 2]. Available from: https://www.cdc.gov/vhf/ebola/history/chronology.html

9. Leroy EM, Kumulungui B, Pourrut X, Rouquet P, Hassanin A, Yaba P, et al. Fruit bats as reservoirs of Ebola virus. Nature. 2005;438(7068):575–6.

10. Towner JS, Pourrut X, Albariño CG, Nkogue CN, Bird BH, Grard G, et al. Marburg virus infection detected in a common African bat. PloS One. 2007;2(8):e764.

11. Judson SD, Fischer R, Judson A, Munster VJ. Ecological contexts of index cases and spillover events of different ebolaviruses. PLoS Pathog. 2016;12(8):e1005780.

12. Jones ME, Schuh AJ, Amman BR, Sealy TK, Zaki SR, Nichol ST, et al. Experimental inoculation of Egyptian rousette bats (Rousettus aegyptiacus) with viruses of the Ebolavirus and Marburgvirus genera. Viruses. 2015;7(7):3420–42.

13. Pigott DM, Golding N, Mylne A, Huang Z, Henry AJ, Weiss DJ, et al. Mapping the zoonotic niche of Ebola virus disease in Africa. Elife. 2014;3:e04395.

14. Pigott DM, Millear AI, Earl L, Morozoff C, Han BA, Shearer FM, et al. Updates to the zoonotic niche map of Ebola virus disease in Africa. Elife. 2016;5:e16412.

15. Schmidt JP, Park AW, Kramer AM, Han BA, Alexander LW, Drake JM. Spatiotemporal fluctuations and triggers of Ebola virus spillover. Emerg Infect Dis. 2017;23(3):415.

16. Hranac CR, Marshall JC, Monadjem A, Hayman DT. Predicting Ebola virus disease risk and the role of African bat birthing. Epidemics. 2019;29:100366.

17. Rulli MC, Santini M, Hayman DT, D’Odorico P. The nexus between forest fragmentation in Africa and Ebola virus disease outbreaks. Sci Rep. 2017;7(1):1–8.

18. Olivero J, Fa JE, Real R, Márquez AL, Farfán MA, Vargas JM, et al. Recent loss of closed forests is associated with Ebola virus disease outbreaks. Sci Rep. 2017;7(1):1–9.

19. Wolfe ND, Daszak P, Kilpatrick AM, Burke DS. Bushmeat hunting, deforestation, and prediction of zoonotic disease. Emerg Infect Dis. 2005;11(12):1822.

20. O’shea TJ, Cryan PM, Cunningham AA, Fooks AR, Hayman DT, Luis AD, et al. Bat flight and zoonotic viruses. Emerg Infect Dis. 2014;20(5):741.

21. Brook CE, Dobson AP. Bats as ‘special’reservoirs for emerging zoonotic pathogens. Trends Microbiol. 2015;23(3):172–80.

22. Meyer CFJ, Kalko EKV, Kerth G. Small-Scale Fragmentation Effects on Local Genetic Diversity in Two Phyllostomid Bats with Different Dispersal Abilities in Panama. Biotropica. 2009;41(1):95–102.

23. Markus N, Hall L. Foraging behaviour of the black flying-fox (Pteropus alecto) in the urban landscape of Brisbane, Queensland. Wildl Res. 2004;31(3):345–55.

24. Mcdonald-Madden E, Schreiber ESG, Forsyth DM, Choquenot D, Clancy TF. Factors affecting Grey-headed Flying-fox (Pteropus poliocephalus: Pteropodidae) foraging in the Melbourne metropolitan area, Australia. Austral Ecol. 2005;30(5):600–8.

25. Castro IJ, Michalski F. Effects of logging on bats in tropical forests. Nat Conserv. 2014 Jul 1;12(2):99–105.

26. Pahari K, Murai S. Modelling for prediction of global deforestation based on the growth of human population. ISPRS J Photogramm Remote Sens. 1999;54(5–6):317–24.

27. Redding DW, Atkinson PM, Cunningham AA, Lo Iacono G, Moses LM, Wood JL, et al. Impacts of environmental and socio-economic factors on emergence and epidemic potential of Ebola in Africa. Nat Commun. 2019;10(1):4531.

28. Fa JE, Olivero J, Farfán MÁ, Márquez AL, Duarte J, Nackoney J, et al. Correlates of bushmeat in markets and depletion of wildlife. Conserv Biol. 2015;29(3):805–15.

29. Bachorec E, Horáček I, Hulva P, Konečnỳ A, Lučan RK, Jedlička P, et al. Spatial networks differ when food supply changes: Foraging strategy of Egyptian fruit bats. PloS One. 2020;15(2):e0229110.

30. Hassanin A, Nesi N, Marin J, Kadjo B, Pourrut X, Leroy É, et al. Comparative phylogeography of African fruit bats (Chiroptera, Pteropodidae) provide new insights into the outbreak of Ebola virus disease in West Africa, 2014–2016. C R Biol. 2016;339(11–12):517–28.

31. Breed AC, Field HE, Smith CS, Edmonston J, Meers J. Bats without borders: long-distance movements and implications for disease risk management. EcoHealth. 2010;7(2):204–12.

32. Richter HV, Cumming GS. First application of satellite telemetry to track African straw-coloured fruit bat migration. J Zool. 2008;275(2):172–6.

33. Stark DJ, Fornace KM, Brock PM, Abidin TR, Gilhooly L, Jalius C, et al. Long-tailed macaque response to deforestation in a Plasmodium knowlesi-endemic area. EcoHealth. 2019;16(4):638–46.

34. Laurance WF, Croes BM, Tchignoumba L, Lahm SA, Alonso A, Lee ME, et al. Impacts of roads and hunting on central African rainforest mammals. Conserv Biol. 2006;20(4):1251–61.

35. Hansen MC, Potapov PV, Moore R, Hancher M, Turubanova SA, Tyukavina A, et al. High-resolution global maps of 21st-century forest cover change. science. 2013;342(6160):850–3.

36. Riitters K, Wickham J, O’Neill R, Jones B, Smith E. Global-scale patterns of forest fragmentation. Conserv Ecol. 2000;4(2).

37. Sulla-Menashe D, Friedl M. MCD12Q1 v006: MODIS/ Terra1Aqua Land Cover Type Yearly L3 Global 500 m SIN Grid. 2019 [cited 2023 Apr 7]; Available from: https://lpdaac.usgs.gov/products/mcd12q1v006/

38. Fick SE, Hijmans RJ. WorldClim 2: new 1-km spatial resolution climate surfaces for global land areas. Int J Climatol. 2017 Oct;37(12):4302–15.

39. Trabucco A, Zomer RJ. Global aridity index and potential evapotranspiration (ET0) climate database v2. CGIAR Consort Spat Inf. 2018;10:m9.

40. Amman BR, Carroll SA, Reed ZD, Sealy TK, Balinandi S, Swanepoel R, et al. Seasonal pulses of Marburg virus circulation in juvenile Rousettus aegyptiacus bats coincide with periods of increased risk of human infection. 2012;

41. Gillespie TR, Jones KE, Dobson AP, Clennon JA, Pascual M. COVID-Clarity demands unification of health and environmental policy. Glob Change Biol. 2021;27(7):1319–21.

42. Aziz SA, Olival KJ, Bumrungsri S, Richards GC, Racey PA. The conflict between pteropodid bats and fruit growers: species, legislation and mitigation. Bats Anthr Conserv Bats Chang World. 2016;377–426.

43. Keita AK, Koundouno FR, Faye M, Düx A, Hinzmann J, Diallo H, et al. Resurgence of Ebola virus in 2021 in Guinea suggests a new paradigm for outbreaks. Nature. 2021;597(7877):539–43.

44. Sims K, Reith A, Bright E, Kaufman J, Pyle J, Epting J, et al. LandScan Global 2022 [Internet]. 2022nd ed. Oak Ridge, TN: Oak Ridge National Laboratory; 2023. Available from: landscan.ornl.gov

45. Golding N, Shearer F. seegSDM: Streamlined Functions for Species Distribution Modelling in the SEEG Research Group. 2022.

46. R Core Team. R: A Language and Environment for Statistical Computing [Internet]. Vienna, Austria: R Foundation for Statistical Computing; 2022. Available from: https://www.R-project.org/

47. Elith J, Leathwick JR, Hastie T. A working guide to boosted regression trees. J Anim Ecol. 2008;77(4):802– 13.

